# Prenatal exposure to environmental contaminants is associated with altered cord serum metabolite profiles in future immune-mediated diseases

**DOI:** 10.1101/2023.11.10.23298353

**Authors:** Bagavathy Shanmugam Karthikeyan, Tuulia Hyötyläinen, Tannaz Ghaffarzadegan, Eric Triplett, Matej Orešič, Johnny Ludvigsson

**Author notes:** Correspondence: Matej Orešič; School of Science and Technology, Örebro University, 702 81 Örebro, Sweden. (editorial correspondence).

## Abstract

Prenatal exposure to environmental contaminants is a significant health concern because it has the potential to interfere with host metabolism, leading to adverse health effects in early childhood and later in life. Growing evidence suggests that genetic and environmental factors, as well as their interactions, play a significant role in the development of autoimmune diseases. In this study, we hypothesized that prenatal exposure to environmental contaminants impacts cord serum metabolome and contributes to the development of autoimmune diseases. We selected cord serum samples from All Babies in Southeast Sweden (ABIS) general population cohort, from infants who later developed one or more autoimmune-mediated and inflammatory diseases: celiac disease (CD), Crohn’s disease (IBD), hypothyroidism (HT), juvenile idiopathic arthritis (JIA), and type 1 diabetes (T1D) (all cases, N = 62), along with matched controls (N = 268). Using integrated exposomics and metabolomics mass spectrometry (MS) based platforms, we determined the levels of contaminants and metabolites. Differences in exposure levels were found between the controls and those who later developed various diseases. High contaminant exposure levels were associated with changes in metabolome, including amino acids and free fatty acids. Specifically, we identified marked associations between metabolite levels and exposure levels of deoxynivalenol (DON), bisphenol S (BPS), and specific per- and polyfluorinated substances (PFAS). Our study suggests that prenatal exposure to specific environmental contaminants alters the cord serum metabolomes, which, in turn, might increase the risk of various immune-mediated disease later in life.

## Introduction

Exposure to environmental contaminants contributes to the global burden of many chronic diseases (Chew et al. 2023; Cui et al. 2016; Diseases and Injuries 2020; Landrigan et al. 2016; Shaffer et al. 2019). Over the past few decades, the prevalence of autoimmune diseases increased in both developed and developing countries, resulting in a high disease burden (Berhan et al. 2011; Carstensen et al. 2020; Eaton et al. 2007; Harjutsalo et al. 2013; Patterson et al. 2009). Autoimmune diseases are often manifested in early childhood and are also common among the pregnant mothers (Eaton et al. 2007; Tincani et al. 2016). They are chronic, impact child growth and development and require long-term management and care (Rosenblum et al. 2012; Wilson et al. 2016). Many studies suggest that a combination of genetic predisposition, environmental and maternal factors as well as their interactions play a significant role in the etiology of autoimmune diseases (Ellis et al. 2014; Oresic et al. 2013; Oresic et al. 2008; Rewers and Ludvigsson 2016; Sen et al. 2019; Sen et al. 2020; Sinisalu et al. 2020; Virolainen et al. 2023).

Exposure of humans to environmental chemicals begins already during the “sensitive window” of human early development, including the prenatal stage (Buhimschi and Buhimschi 2012; Karthikeyan et al. 2021; Landrigan and Goldman 2011; Robinson and Vrijheid 2015). Prenatal exposure to PFAS and other contaminants have been associated with abnormal metabolism and later progression to autoimmune diseases such as T1D (McGlinchey et al. 2020), CD (Sen et al. 2019; Sinisalu et al. 2020), IBD (Filimoniuk et al. 2020) later. PFAS exposure, for instance, alters the levels of phospholipids and contributes to the risk of T1D (McGlinchey et al. 2020). Although most autoimmune diseases share common pathogenicity and genetic risk factors (Ilonen et al. 2016; Sen et al. 2019), their underlying pathogenic mechanisms are poorly understood. Beside exposure to environmental chemicals, perinatal factors such as low birth weight (Katsarou et al. 2017), the gut microbiome (Belteky et al. 2023; Khan and Wang 2019; Kindgren et al. 2023; Kostic et al. 2015; Vatanen et al. 2018; Weis 2018) and maternal diet (Johnson et al. 2021; Johnson et al. 2019; Virtanen et al. 2012) are also attributed to the progression of autoimmune diseases.

Given the potential impact of exposure to environmental contaminants and the role of maternal factors in the progression of autoimmune diseases (Hyotylainen et al. 2023), it is important to characterize the prenatal and early-life exposome to better understand the pathogenesis of autoimmune diseases. Herein, we hypothesized that exposure to environmental contaminants impacts cord serum metabolome, which may contribute to the development of one or more autoimmune diseases in the general population cohort (All Babies In Southeast Sweden, ABIS) (Ludvigsson et al. 2001; Nygren et al. 2015). We quantified levels of contaminants and metabolite profiles from cord serum collected at birth, using integrated exposomics and metabolomics approaches. We investigated (i) the levels of exposure and significant differences between controls and cases, (ii) associations of contaminant exposure with cord serum metabolic profiles, and (iii) the impact of contaminant exposure levels on cord serum metabolic profiles.

## Materials and methods

### Study design

ABIS, a general population cohort consists of 17000 children born 1^st^ of Oct 1997-1^st^ of Oct 1999, followed prospectively with regular follow-ups. ABIS is connected to the Swedish National Diagnosis Register which give information about diagnosis of autoimmune disease. Stool samples were collected from ca 1800 individuals at 1 year of age, and microbiome studies have been performed (Belteky et al. 2023; Hyotylainen et al. 2023). The present study includes subjects (N=62) from this group who later developed one or more autoimmune and inflammatory diseases such as cCeliac disease (CD), Crohns disease (IBD), hypothyroidism (HT), juvenile idiopathic arthritis (JIA) and type1 diabetes (T1D) along with their matching controls (N=268). The cord blood samples collected during birth were subjected to metabolomics analysis. **Fig. 1** summarizes the study design and integrated workflow.

**Fig. 1.**
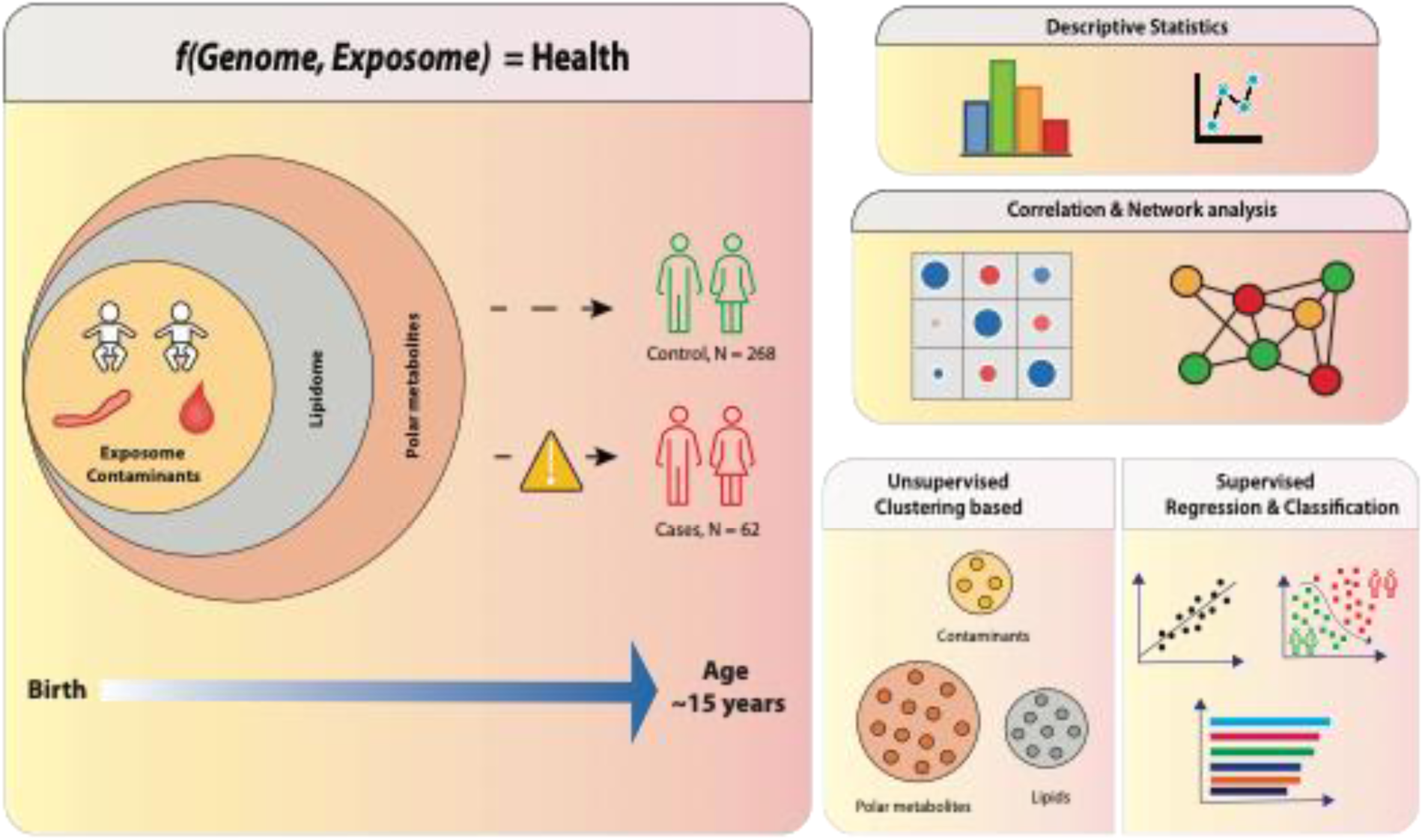
Summary of our work, which aimed to investigate how prenatal exposure to environmental contaminants alters the cord serum metabolome in the ABIS cohort. We used metabolomics to determine the levels of exposure to environmental contaminants and metabolites in the cord blood. Our work involved three main stages. Firstly, we examined the levels of exposure and significant differences between control (N = 268) and cases (N = 62). Secondly, we studied the associations of contaminant exposure with cord serum metabolic profiles. Finally, we investigated the impact of contaminant exposure on cord serum metabolites. Overall, our work sheds light on the effects of environmental contaminants on the cord serum metabolome, which may have implications for the future progression of autoimmune diseases.

This study was performed in accordance with the Declaration of Helsinki. The ABIS study was approved by the Research Ethics Committees of the Faculty of Health Sciences at Linköping University, Sweden, 1997/96287 and 2003/03-092 and the Medical Faculty of Lund University, Sweden (DNR 99227, DNR 99321). All participating parents gave their informed consent to participate in ABIS after oral, written and video information. ABIS connection to national register approved by the Research Ethics Committees of the Faculty of Health Sciences at Linköping University, Sweden, DNR 05-513, and 2018/380-32.

### Analysis of metabolome and environmental contaminants

A total of 360 cord blood samples were randomized and analyzed as described below. Shortly, two methods were applied for separate extraction of lipids and polar/semipolar metabolites and the extracts were then analyzed using an ultra-high-performance liquid chromatography quadrupole time-of-flight mass spectrometry (UHPLC-QTOFMS) as described previously (Hyotylainen et al. 2023) and the data were processed using MZmine 2.53 (Pluskal et al. 2010). Quantification was performed using calibration curves and the identification was done with a custom data base, with identification levels 1 and 2 (Metabolomics Standards Initiative). Quality control was performed by analyzing pooled quality control samples. In addition, extracted blank samples, standards compounds, and reference plasma (NIST SRM 1950); purchased from the National Institute of Standards and Technology at the US Department of Commerce (Washington, DC, USA)) were analyzed as part of the quality control procedure.

#### Analysis of molecular lipids

10 µl of serum was mixed with 10 µl 0.9% NaCl and extracted with 120 µl of CHCl_3_: MeOH (2:1, v/v) solvent mixture containing internal standard mixture (c = 2.5 µg/ml; 1,2-diheptadecanoyl-sn-glycero-3-phosphoethanolamine (PE(17:0/17:0)), N-heptadecanoyl-D-erythro-sphingosylphosphorylcholine (SM(d18:1/17:0)), N-heptadecanoyl-D-erythro-sphingosine (Cer(d18:1/17:0)), 1,2-diheptadecanoyl-sn-glycero-3-phosphocholine (PC(17:0/17:0)), 1-heptadecanoyl-2-hydroxy-sn-glycero-3-phosphocholine (LPC(17:0)) and 1-palmitoyl-d31-2-oleoyl-sn-glycero-3-phosphocholine (PC(16:0/d31/18:1)) and, triheptadecanoylglycerol (TG(17:0/17:0/17:0)). The samples were vortexed and let stand on the ice for 30 min before centrifugation (9400 rcf, 3 min). 60 µl of the lower layer of was collected and diluted with 60 µl of CHCl_3_: MeOH. The samples were kept at −80 °C until analysis.

Samples were analyzed by UHPLC-QTOFMS (Agilent Technologies; Santa Clara, CA, USA). The analysis was carried out on an ACQUITY UPLC BEH C18 column (2.1 mm × 100 mm, particle size 1.7 μm) by Waters (Milford, USA). he eluent system consisted of (A) 10 mM NH_4_Ac in H_2_O and 0.1% formic acid and (B) 10 mM NH_4_Ac in ACN: IPA (1:1) and 0.1% formic acid. The gradient was as follows: 0-2 min, 35% solvent B; 2-7 min, 80% solvent B; 7-14 min 100% solvent B. The flow rate was 0.4 ml/min.

The following steps were applied in data processing with MZmine 2.53: (i) Mass detection with a noise level of 1000, (ii) Chromatogram builder with a minimum time span of 0.08 min, minimum height of 1000 and a m/z tolerance of 0.006 m/z or 10.0 ppm, (iii) Chromatogram deconvolution using the local minimum search algorithm with a 70% chromatographic threshold, 0.05 min minimum RT range, 5% minimum relative height, 1200 minimum absolute height, a minimum ration of peak top/edge of 1.2 and a peak duration range of 0.08–5.0, (iv), Isotopic peak grouper with a m/z tolerance of 5.0 ppm, RT tolerance of 0.05 min, maximum charge of 2 and with the most intense isotope set as the representative isotope, (v) Join aligner with a m/z tolerance of 0.009 or 10.0 ppm and a weight for of 2, a RT tolerance of 0.15 min and a weight of 1 and with no requirement of charge state or ID and no comparison of isotope pattern, (vi) Peak list row filter with a minimum of 10% of the samples (vii) Gap filling using the same RT and m/z range gap filler algorithm with an m/z tolerance of 0.009 m/z or 11.0 ppm, (vii) Identification of lipids using a custom database search with an m/z tolerance of 0.008 m/z or 8.0 ppm and a RT tolerance of 0.25 min. Identification of lipids was based on an in-house librarybased on LC-MS/MS data on retention time and mass spectra. The identification was done with a custom data base, with identification levels 1 and 2, *i.e.*, based on authentic standard compounds (level 1) or based on MS/MS identification (level 2).

Quantification of lipids was performed using a 7-point internal calibration curve (0.1-5 µg/mL) using the following lipid-class specific authentic standards: using 1-hexadecyl-2-(9Z-octadecenoyl)-sn-glycero-3-phosphocholine (PC(16:0e/18:1(9Z))), 1-(1Z-octadecenyl)-2-(9Z-octadecenoyl)-sn-glycero-3-phosphocholine (PC(18:0p/18:1(9Z))), 1-stearoyl-2-hydroxy-sn-glycero-3-phosphocholine (LPC(18:0)), 1-oleoyl-2-hydroxy-sn-glycero-3-phosphocholine (LPC(18:1)), 1-palmitoyl-2-oleoyl-sn-glycero-3-phosphoethanolamine (PE(16:0/18:1)), 1-(1Z-octadecenyl)-2-docosahexaenoyl-sn-glycero-3-phosphocholine (PC(18:0p/22:6)) and 1-stearoyl-2-linoleoyl-sn-glycerol (DG(18:0/18:2)), 1-(9Z-octadecenoyl)-sn-glycero-3-phosphoethanolamine (LPE(18:1)), N-(9Z-octadecenoyl)-sphinganine (Cer(d18:0/18:1(9Z))), 1-hexadecyl-2-(9Z-octadecenoyl)-sn-glycero-3-phosphoethanolamine (PE(16:0/18:1)) from Avanti Polar Lipids, 1-Palmitoyl-2-Hydroxy-sn-Glycero-3-Phosphatidylcholine (LPC(16:0)), 1,2,3 trihexadecanoalglycerol (TG(16:0/16:0/16:0)), 1,2,3-trioctadecanoylglycerol (TG(18:0/18:0/18:)) and 3β-hydroxy-5-cholestene-3-stearate (ChoE(18:0)), 3β-Hydroxy-5-cholestene-3-linoleate (ChoE(18:2)) from Larodan, were prepared to the following concentration levels: 100, 500, 1000, 1500, 2000 and 2500 ng/mL (in CHCl3:MeOH, 2:1, v/v) including 1250 ng/mL of each internal standard.

#### Analysis of polar metabolites

40 µl of serum sample was mixed with 90 µl of cold MeOH/H2O (1:1, v/v) containing the internal standard mixture (Valine-d8, Glutamic acid-d5, Succinic acid-d4, Heptadecanoic acid, Lactic acid-d3, Citric acid-d4. 3-Hydroxybutyric acid-d4, Arginine-d7, Tryptophan-d5, Glutamine-d5, each at at c= 1 µgmL-1 and 1-D4-CA,1-D4-CDCA,1-D4-CDCA,1-D4-GCA,1-D4-GCDCA,1-D4-GLCA,1-D4-GUDCA,1-D4-LCA,1-D4-TCA, 1-D4-UDCA, each at 0.2 1 µgmL-1) for protein precipitation. The tube was vortexed and ultrasonicated for 3 min, followed by centrifugation (10000 rpm, 5 min). After centrifuging, 90 µl of the upper layer of the solution was transferred to the LC vial and evaporated under the nitrogen gas to the dryness. After drying, the sample was reconstituted into 60 µl of MeOH: H_2_O (70:30).

Analyses were performed on an Agilent 1290 Infinity LC system coupled with 6545 QTOFMS interfaced with a dual jet stream electrospray (dual ESI) ion source (Agilent Technologies; Santa Clara, CA, USA) was used for the analysis. Aliquots of 10 μL of samples were injected into the Acquity UPLC BEH C18 2.1 mm × 100 mm, 1.7-μm column (Waters Corporation, Wexford, Ireland), fitted with a C18 precolumn (Waters Corporation, Wexford, Ireland). The mobile phases consisted of (A) 2 mM NH_4_Ac in H_2_O: MeOH (7:3) and (B) 2 mM NH_4_Ac in MeOH. The flow rate was set at 0.4 mLmin-1 with the elution gradient as follows: 0-1.5 min, mobile phase B was increased from 5% to 30%; 1.5-4.5 min, mobile phase B increased to 70%; 4.5-7.5 min, mobile phase B increased to 100% and held for 5.5 min. A post-time of 5 min was used to regain the initial conditions for the next analysis. The total run time per sample was 20 min. The dual ESI ionization source was settings were as follows: capillary voltage was 4.5 kV, nozzle voltage 1500 V, N2 pressure in the nebulized was 21 psi and the N2 flow rate and temperature as sheath gas was 11 Lmin-1 and 379 °C, respectively. In order to obtain accurate mass spectra in MS scan, the m/z range was set to 100-1700 in negative ion mode. MassHunter B.06.01 software (Agilent Technologies; Santa Clara, CA, USA) was used for all data acquisition.

#### MS data processing was performed using same parameters as in lipidomic analysis

Quantitation was done using 6-point calibration (PFOA c= 3.75-120 ng/mL, bile acids c= 20-640 ng/mL, polar metabolites c=0.1 to 80 μg/mL). Quantification of other bile acids was done using the following compounds: chenodeoxycholic acid (CDCA), cholic acid (CA), deoxycholic acid (DCA), glycochenodeoxycholic acid (GCDCA), glycocholic acid (GCA), glycodehydrocholic acid (GDCA), glycodeoxycholic acid (GDCA), glycohyocholic acid (GHCA), glycohyodeoxycholic acid (GHDCA), glycolitocholic acid (GLCA), glycoursodeoxycholic acid (GUDCA), hyocholic acid (HCA), hyodeoxycholic acid (HDCA), litocholic acid (LCA), alpha-muricholic acid (αMCA), tauro-alpha-muricholic acid (T-α-MCA), tauro-beta-muricholic acid(T-β-MCA), taurochenodeoxycholic acid (TCDCA), taurocholic acid (TCA), taurodehydrocholic acid (THCA), taurodeoxycholic acid (TDCA), taurohyodeoxycholic acid (THDCA), taurolitocholic acid (TLCA), tauro-omega-muricholic acid (TωMCA) and tauroursodeoxycholic acid (TDCA) and polar metabolites was done using alanine, citric acid, fumaric acid, glutamic acid, glycine, lactic acid, malic acid, 2-hydroxybutyric acid, 3-hydroxybutyric acid, linoleic acid, oleic acid, palmitic acid, stearic acid, cholesterol, fructose, glutamine, indole-3-propionic acid, isoleucine, leucine, proline, succinic acid, valine, asparagine, aspartic acid, arachidonic acid, glycerol-3-phosphate, lysine, methionine, ornithine, phenylalanine, serine and threonine.

#### QC/QA

Quality control was accomplished both for lipidomics, polar metabolites and PFAS analysis by including blanks, pure standard samples, extracted standard samples, pooled quality control samples and standard reference plasma samples (NIST SRM 1950). The pooled sample were prepared by taking an aliqout (10 µl) of each extract, separately for lipidomic and polar metabolite methods, then pooling them, and aliquoting the pool into separate vials. In lipidomic and metabolomic analyses, lipids that had >30% RSD in the pooled QC samples (an equal aliquot of each sample pooled together) or that were present at high concentrations in the extracted blank samples (ratio between samples to blanks < 5) were excluded from the data analyses.

### Statistical analysis

#### Data pre-processing and clustering

In this study, all data analyses were conducted using the R statistical programming language (version 4.1.2) (https://www.r-project.org/). The exposure datasets were pre-processed by log2 transformation and scaling to zero mean and unit variance (auto-scaled). For contaminant exposure analyses, individual contaminant and cluster-level analyses were performed. To cluster the contaminant data, we utilized the ‘*mclust*’ R package (version 5.4.10) for model-based clustering, selecting the model type and the number of clusters based on the highest Bayesian Information Criterion (BIC). To better understand the impact of exposure, we also incorporated lipidomics and metabolomics data from our previous study (Hyotylainen et al. 2023), which included eight lipid clusters (LCs) and twelve polar metabolite clusters (PCs), along with their individual features.

#### Demographic data and covariates

In terms of demographic data and covariates, the median age at the time of diagnosis for subjects who later developed autoimmune diseases was 15 years. We obtained information on birthweight, maternal age, gestational age, and BMI from the questionnaire. Additionally, we utilized birthweight and gestational age to calculate birthweight for gestational age (BWGA) Z-score, utilizing internationally validated infant growth charts developed by Fenton (Chou et al. 2020; Fenton and Kim 2013).

#### Correlation and partial correlation analysis

Pairwise Spearman’s correlation between contaminants, lipid clusters (LCs), Polar metabolite clusters (PCs) and demographic variables (Z-score, Maternal age, BMI) was calculated and visualized using ‘*corrplot*’ R package (version 0.92). Two correlation plots were generated separately for control and cases. The correlation between variables visualised in the form of a matrix plot refers to positive and negative correlations and the strength of the association is referred to by the size of the dot or filled circles.

The Debiased Sparse Partial Correlation algorithm (DSPC) (Basu et al. 2017) was used to estimate partial correlation networks and visualized in the form of a chord diagram using ‘*circlize*’, R package (version 0.4.15) with edge ranges between ±0.14 to 1.0 and showing only correlations across contaminants, LCs, PCs and demographic variables.

#### Univariate statistical analysis

To understand the impact of contaminant exposure levels on cord serum metabolome, the subjects were assigned to four quartiles based on the exposure levels. A two-way analysis of variance (ANOVA) test was performed followed by post-hoc Tukey’s test by using quartiles (Q1 to Q4) and subjects (cases and control) as factor variables. ANOVA test helps to identify any significant changes in the lipid or metabolite clusters and post-hoc Tukey’s test helps to identify the specific quartiles between which significant changes are observed.

#### Regression and classification analysis

Predictive logistic ridge regression (LRR) was performed to investigate the impact of individual contaminants on the stratification of autoimmune cases and controls. We have adapted the L2 regularization strategy to avoid multicollinearity among highly correlated predictors. Regularized regression modelling was performed using the ‘*glmnet*’ package in R (version 4.1-4). The hyper-parameter λ_minimum_ was determined by 10-fold cross-validation using the ‘*cv.glmnet*’ function from ‘*glmnet*’. The models were adjusted for Z-score, Maternal age and BMI. The accuracy of prediction was determined by AUCs, where the mean AUC of the model was estimated by bootstrapping, by resampling the exposure dataset into training (80%) and testing (20%) 10,000 times. All LRR models with a threshold of AUC > 0.60 were considered. Downsampling was performed to address the class imbalance problem (cases, n=62, controls, n >62). The ‘*caret*’ package (version 4.1.3) was used for the partition of data and the best models (based on mean AUCs) were assessed using Receiver Operating Characterisitic (ROC) curves using the ‘*ROCR*’ package. Additionally, we have performed a stepwise recursive feature elimination scheme to identify the minimum number of predictors that are needed to maximize the outcome.

To investigate the effect of contaminant exposure on the cord blood metabolome, we employed linear regression with L2 regularization (LR), using individual contaminant concentrations as predictors and the concentrations of significantly altered cord blood lipid or polar metabolites (and their cluster) as the response variable. The hyper-parameter λ_minimum_, which corresponds to the minimum cross-validation error, was selected through 10-fold cross-validation. We partitioned the data and performed resampling (10,000 iterations) as described earlier. The mean R square was used to estimate the accuracy of prediction and the significant impact of contaminant exposure on the cord blood metabolome.

Additionally, we determined the ranks of the predictors using LR and LRR modelling. For the LRR models, the ranks of the predictors were estimated based on the unit absolute differences in the odds ratio, while for the LR models, the ranks were based on the ridge coefficients normalized with the maximum value.

#### Pathway analysis

Pathway enrichment analysis comparing cases versus controls for Deoxynivalenol (DON) impact polar metabolites was performed using the MetaboAnalyst 5.0 web platform with the Functional Analysis (MS Peaks) module (Pang et al. 2022). The input data for the pathway analysis consisted of complete high-resolution LC-MS spectral peak data obtained in negative ionization mode with a mass tolerance of 10 ppm. Linear regression analysis was performed to estimate the association between DON and polar metabolites while adjusting for Z-score, Maternal age, and BMI. The whole input peak list with FDR-corrected p-values and T-score was used for the pathway analysis. Overrepresented pathways were estimated against the background human scale metabolic model MNF (from MetaboAnalyst Mummichog package) and Kyoto Encyclopedia of Genes and Genomes (KEGG) pathways for Homo sapiens to determine the relative significance of the identified pathways (Li et al. 2020). The MetaboAnalyst 5.0 metabolomics pathway analysis (MetPA) tool(Xia and Wishart 2010) was used to calculate the Pathway Impact Scores (Chong et al. 2018; Pang et al. 2022).

## Results

### Metabolomic analysis of the cord blood

**Fig. 1** summarizes the integration of exposomics and metabolomics workflows in the ABIS cohort. Cord serum samples were analyzed for a total of 545 lipids and 3417 polar metabolites, which were further grouped into 8 lipid clusters and 12 polar metabolite clusters, respectively. We previously found significant associations between the metabolite clusters and demographic variables or clinical parameters such as gestational age, maternal age, and birth weight (Hyotylainen et al. 2023). To account for these associations, we used the Z-score as calculated from birth weight, gestational age, and maternal BMI as covariates in our analysis.

### Levels of contaminants in the cord blood

A total of 20 contaminants, including several PFAS compounds, were detected in cord blood samples from both control and case groups (**Table S1**). Differences (p<0.05) in concentration levels between control and case groups were observed for Perfluorooctanoic acid Branched 2, Environmental Contaminant 1, Perfluorooctanoic acid Linear 1, Perfluorooctanoic acid Linear 2 and Methylparaben (**Fig. 2**). At the individual disease level, Environmental Contaminant 1, Perfluorooctanoic acid Linear 2 and Perfluorooctanoic acid Branched 2 showed differences (p<0.05) in concentration levels between control and individual disease groups (**Fig. S1**). The contaminants were reduced to four clusters (CC1-CC4) consisting of eight contaminants, including Bisphenol S, Deoxynivalenol, Monobutyl phthalate, and a-Zearalanol in CC1; Ethylparaben, Methylparaben, and Propylparaben in CC2; Perfluorohexanesulfonic acid (PFHxS) and Perfluorohexanesulfonic acid Branched (PFHxSBr) in CC3; and seven PFAS and their fragments as part of CC4 (**Table S1**).

**Fig. 2.**
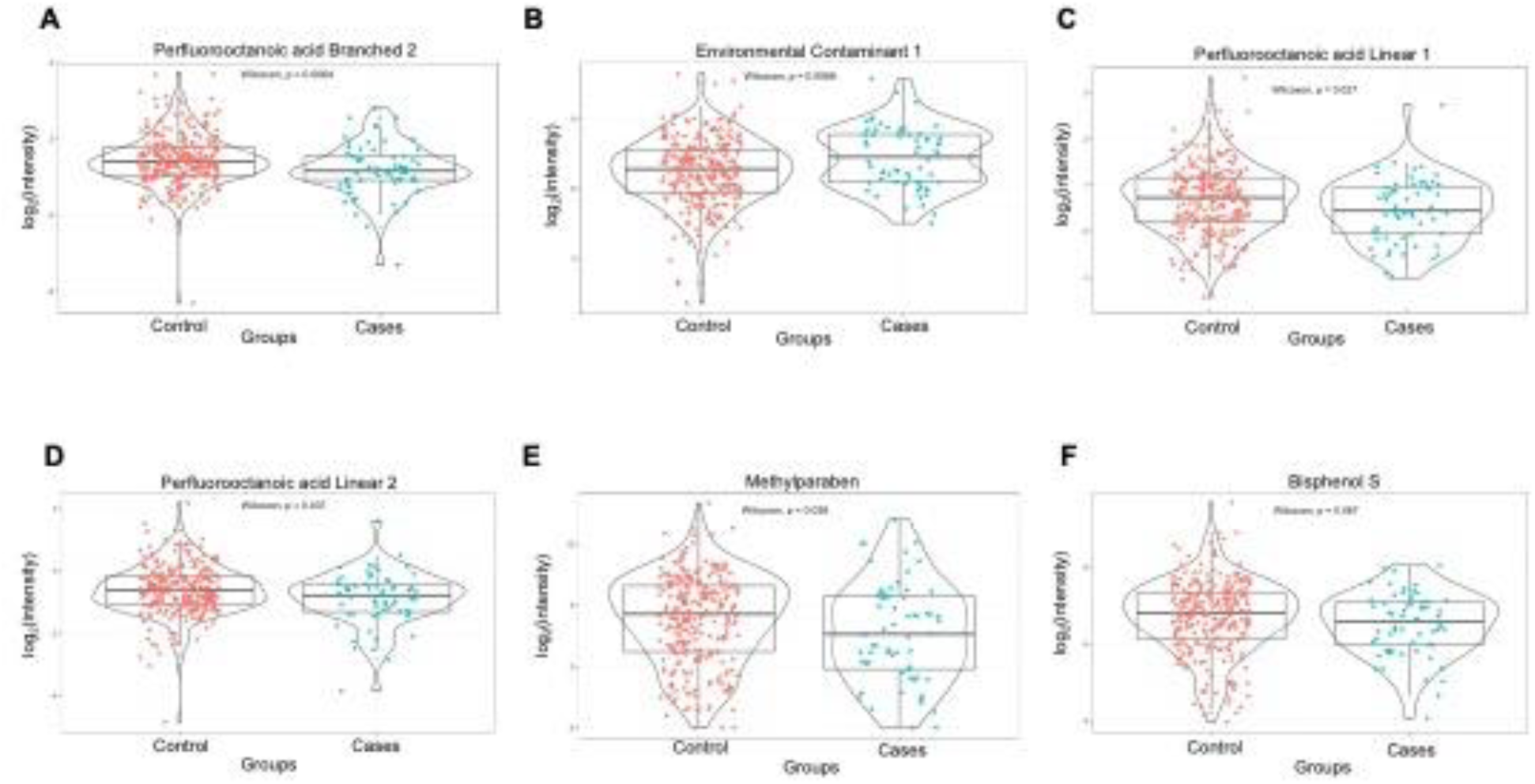
Box plots that illustrate the levels of selected contaminants in control and cases. The violin plots (A-F) on top of the box plots depict the distribution of the selected contaminants (log2 intensities). To test the mean difference between the control and cases, we conducted a Wilcoxon test. The p-values are provided to indicate the significance levels for the mean differences between the two groups for each contaminant (A-F). Specifically, p < 0.05 indicates statistical significance, and p < 0.1 suggests a trend toward significance. Overall, these results help to identify specific contaminants that may contribute to the altered cord serum metabolome in cases.

### Exposure level of contaminants as a predictor for immune-mediated diseases

We employed predictive logistic ridge regression (LRR) models to stratify controls and cases based on their contaminant concentrations. The models were fitted using all predictors or by using the stepwise recursive feature elimination (RFE) method. The mean area under the curve (AUC) values for the models were 0.65 (95% CI 0.63-0.67) when using all predictors and 0.67 (95% CI 0.66-0.68) when using the stepwise RFE method (**Fig. S2**). Our results showed that the contaminant concentration levels have a modest potential to differentiate controls from autoimmune diseases, as indicated by the mean AUC values (**Fig. S2**). The ranks of individual contaminants (predictors) for separating controls and cases were estimated based on the unit absolute difference in odds ratios (**Fig. S2A**).

### Associations between contaminants and cord serum metabolic profiles

We found significant associations between contaminants and cord serum metabolic profiles (**Fig. 3**). Specifically, more associations were observed between PFAS exposures and metabolic profiles in cases than in controls. Maternal age was positively associated with metabolite cluster PC1 in cases but not in controls (**Fig. 3**). We also performed partial correlation network analysis to identify non-spurious associations and **Fig. 4** shows the marked associations between contaminants and cord serum metabolic profiles along with demographic variables. In the case group (**Fig. 4B**), the covariates Z-score, maternal age, and BMI showed a stronger association with exposure and cord serum metabolic profiles compared to the control group (**Fig. 4A**). The mycotoxins including deoxynivalenol were found to be associated with PC2 (phosphatidylcholines) and PC10 (unknowns), while a-zearalanol was associated with PC1 (lysophosphatidylcholines, sphingomyelins, and ceramides) (**Fig. 4**).

**Fig. 3.**
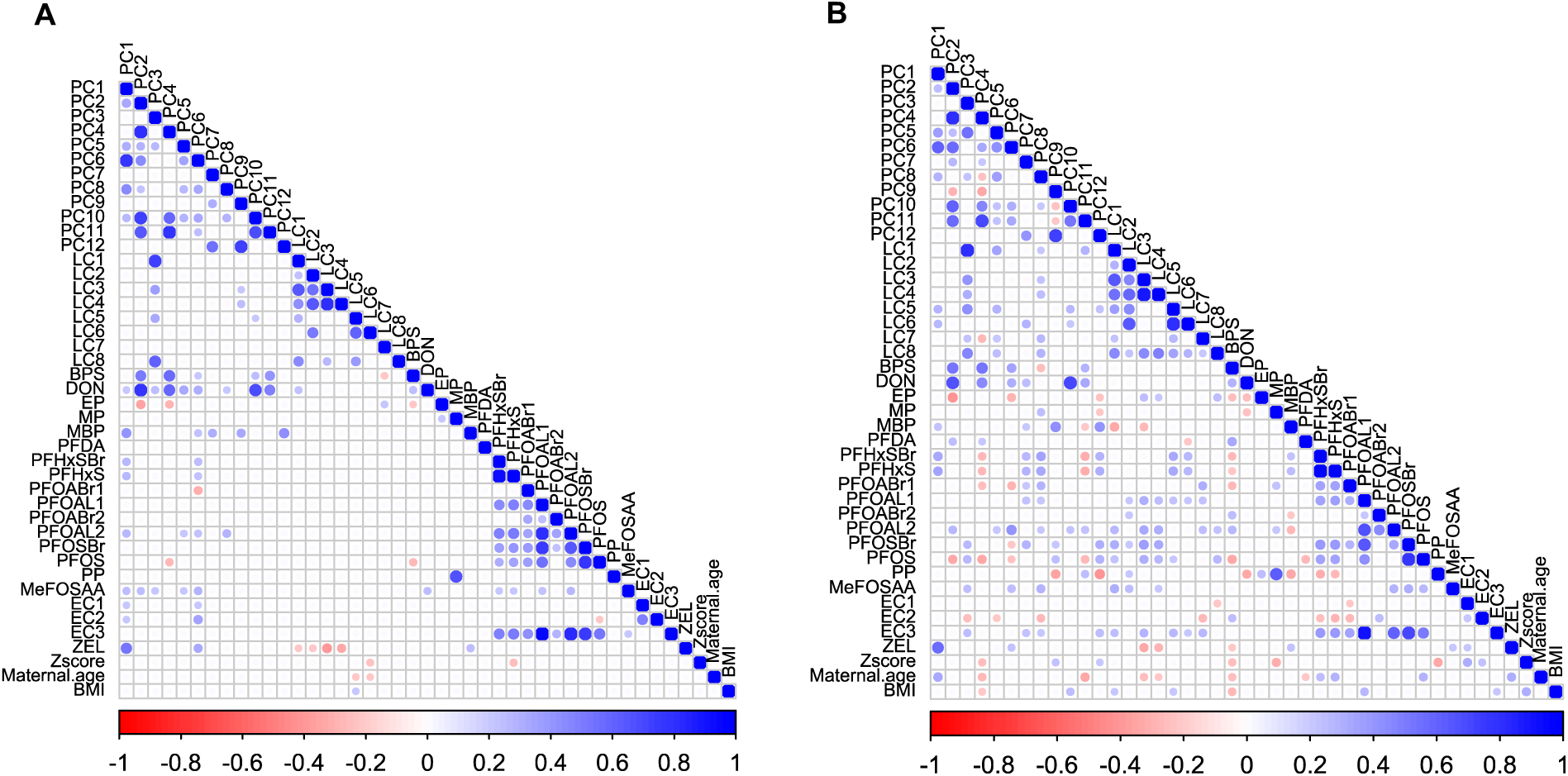
Correlation plots that depict the relationships between contaminant exposure, metabolite clusters, and demographic data in controls (A) and cases (B). We used pairwise Spearman correlation to calculate the correlation coefficients between all cluster variables, contaminants, and demographic variables in the ABIS cohort. Positive and negative correlations are denoted by blue and red colours, respectively. The size of the dot in each cell corresponds to the strength of the pairwise correlation. To improve visualization, we only show correlations between +/− 0.20 to 1.0 in the plots. Overall, these correlation plots provide a comprehensive overview of the complex relationships between environmental contaminants, metabolites, and demographic factors in the ABIS cohort.

**Fig. 4.**
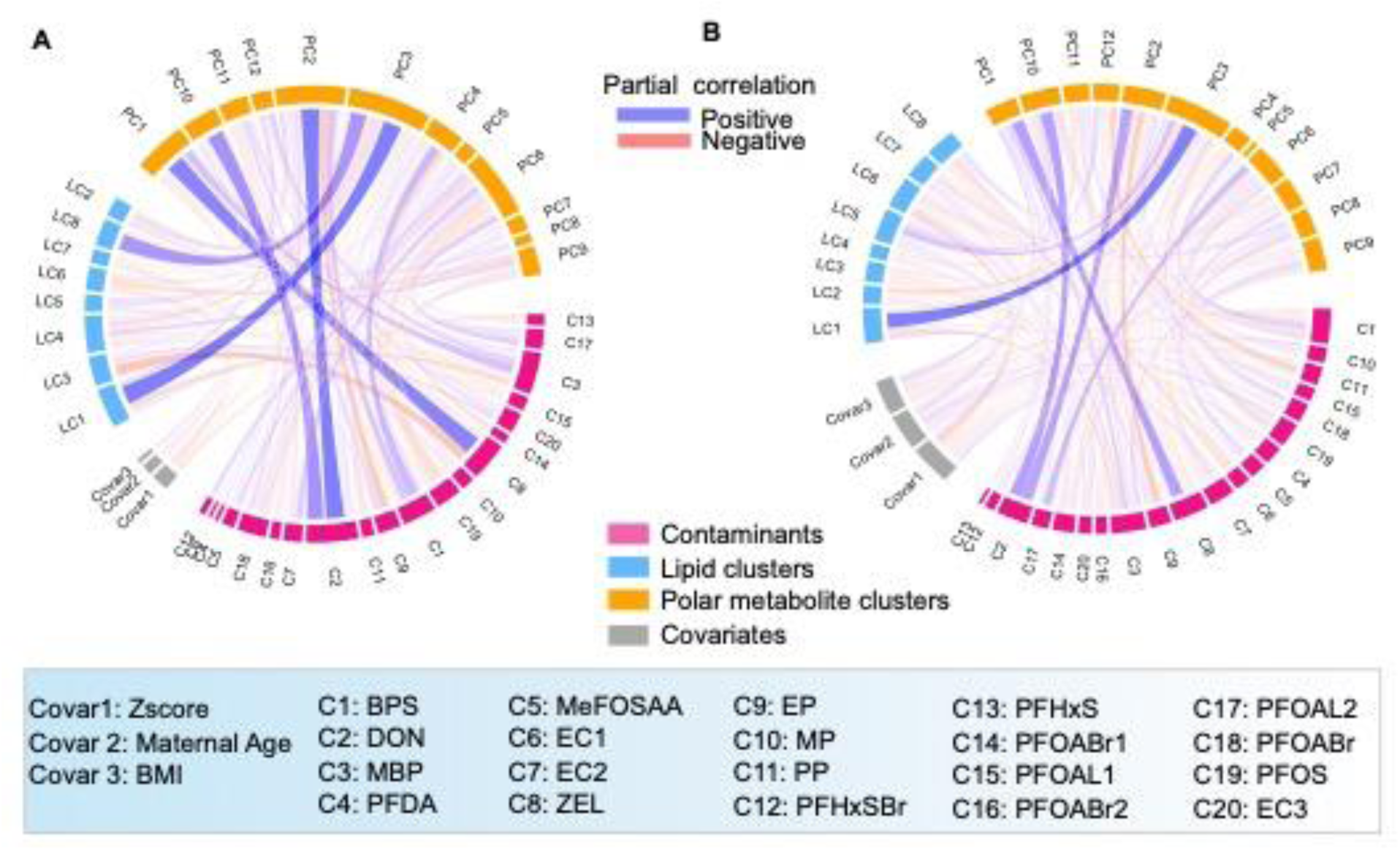
Partial correlation network in the form of a chord diagram that shows the associations between contaminant exposure, metabolite clusters, and demographic data in controls (A) and cases (B). To filter out spurious or indirect correlations between variables, we used the Debiased Sparse Partial Correlation (DSPC) algorithm (Basu et al. 2017) to only show direct correlations. We used a conservative cut-off between +/− 0.14 to 1.0 to visualize the correlations and project only correlations across groups (Contaminants, metabolite clusters, and covariates/demographic data). Positive and negative correlations are denoted by blue and red lines, respectively. Overall, this partial correlation network provides a more detailed view of the complex relationships between environmental contaminants, metabolites, and demographic factors in the ABIS cohort.

### Impact of contaminant exposure on cord serum metabolites associated with immune-mediated diseases

The samples were stratified into quartiles based on their level of exposure to contaminants, and the impact of exposure on metabolite levels was assessed at both individual contaminant levels and cluster levels (CC1-CC4) (**Tables S2-S5**). The polar metabolite clusters displayed more significant mean differences between the highest (Q4) and lowest (Q1) quartiles, as shown in **Table S4-S5**. In CC1, significant mean differences between Q4 and Q1 were observed for LC3, LC4, and LC7 at the lipid cluster level (**Table S3**). In CC3, which includes perfluorohexanesulfonic acid (PFHxS) and branched (PFHxSBr), significant mean differences between the highest and lowest quartiles were observed for LC5 and LC6 (**Table S3** and **Fig. 5H**).

**Fig. 5.**
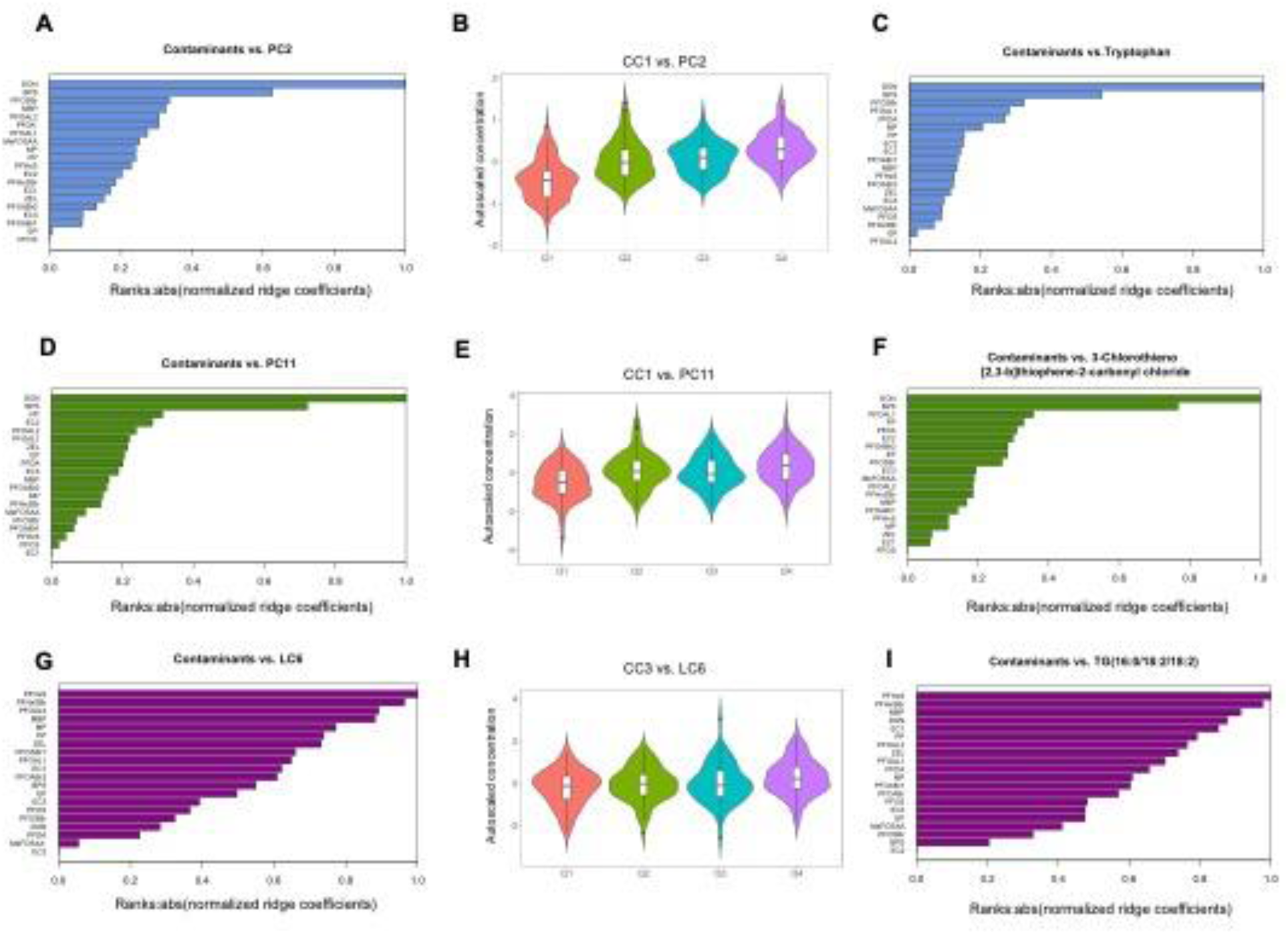
Impact of environmental contaminants on cord serum metabolites. Horizontal bar plots (A, D, G) and (C, F, I) display the ranks of contaminants as predictors of metabolite clusters (PC2, PC11, and LC6) and individual metabolites (Tryptophan, 3-Chlorothieno [2,3-b]thiophene-2-carbonyl chloride, and TG(16:0/18:2/18:2)), respectively. The potential impact of contaminants on metabolite clusters or individual metabolites is determined by their rank at the top of the bar plot. The ranks are based on their absolute normalized (ridge) regression coefficients. Violin plots (B, E, H) show the levels of metabolites (clusters) associated with levels of exposure to contaminants from contaminants clusters 1 (CC1) and 3 (CC3). The violin plot represents the density of the sample within each quartile, and their distribution is represented using a box plot at the centre. Two-way ANOVA followed by post-hoc Tukeýs HSD test was used to compare the mean difference between levels of metabolites (along quartiles).

Linear ridge regression (LR) was performed to determine the quantitative effect of contaminant concentration levels on cord serum metabolic profiles. The results showed that polar metabolites in cord serum were more highly impacted by contaminant exposure than lipid levels. Specifically, six polar metabolite clusters, PC2 (R^2^ = 0.72), PC6 (R^2^ = 0.53), PC4 (R^2^ = 0.52), PC1 (R^2^ = 0.48), PC10 (R^2^ = 0.48), and PC11 (R^2^ = 0.32), showed significant associations with exposure levels (**Fig. 5** and **Fig. S3**). At the individual metabolite level, amino acids such as tryptophan (**Fig. 5C**), Serine of PC2, and 3-Chlorothieno [2,3-b]thiophene-2-carbonyl chloride of PC11 showed a significant impact (**Fig. 5F**). According to the ranks of the predictors (contaminants), deoxynivalenol (DON) and Bisphenol S were the top linear predictors of cord serum metabolites and clusters PC2 and PC11 (**Fig. 5A-F**).

Although the contaminants from clusters CC1 and CC3 showed a significant association between quartiles (Q4 vs. Q1) and lipid cluster levels LC3, LC4, LC5, LC6, and LC7, their strength of association based on LR models was comparatively weaker (**Fig. 5G-I** and **Fig. S3**). For example, the lipid cluster LC6, which mainly comprises triglycerides containing monounsaturated fatty acid (MUFA) and polyunsaturated fatty acids (PUFA), showed a weaker association (R^2^ = 0.04) with contaminant exposures (**Fig. 5G-I**).

### Pathway analysis of deoxynivalenol exposure

Metabolic pathway enrichment analysis was performed to evaluate the impact of DON on polar metabolites in both control and case groups separately. DON was found to be the top predictor that impacted polar metabolite clusters PC2, PC4, PC10, and PC11, as shown in **Fig. 5** and **Fig. S3**. Both Mummichog and GeneSet Enrichment Analysis (GSEA) algorithms were utilized using MetaboAnalyst 5.0 (Li et al. 2020; Pang et al. 2022). Based on the pathways identified by the impact of DON exposure, both control and case groups showed common and specific metabolic pathways, as presented in **Tables S6-S9**.

The MFN pathway map revealed that DON exposure was associated with ‘Tyrosine and Tryptophan metabolism’ in the control group but not in cases. Also ‘Glutathione Metabolism’, ‘Alanine and Aspartate Metabolism’, and ‘Glycerophospholipid metabolism’ were found to be associated with exposure to DON in cases, but not in controls (**Fig. 6A** and **6C**; **Tables S6** and **S8**). Similarly, based on the KEGG pathway maps, ‘Aminoacyl-tRNA biosynthesis’ and ‘Glycine, serine, and threonine metabolism’ were common among control and case groups, while several other metabolic pathways were specific to each group (**Fig. 6B** and **6D**, **Tables S7** and **S9**). In summary, the pathway enrichment analysis provided insights into the metabolic pathways affected by DON exposure in both control and case groups. The results highlight the differences in the impacted pathways between the two groups based on the exposure to DON.

**Fig. 6.**
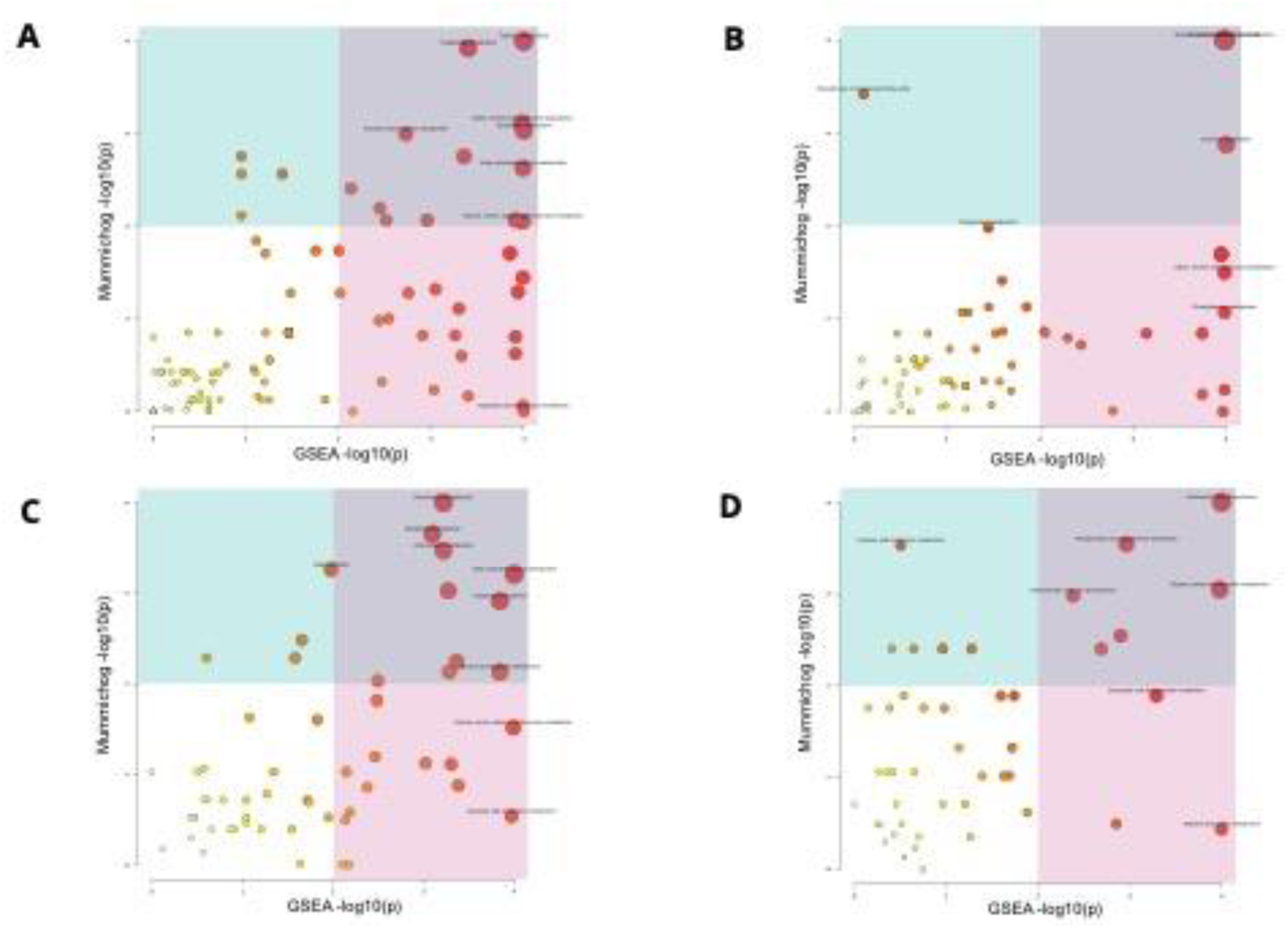
Pathway enrichment analysis comparing cases versus controls for Deoxynivalenol (DON) impact polar metabolites. The scatter plots depict the p-values using two different pathway maps: MFN pathway maps on the left panels and KEGG pathway maps on the right panels. The pathway analysis methods Mummichog and GSEA are used on the y-axis and x-axis, respectively, for both control (A, B) and cases (C, D). The size of the circle on each scatter plot represents the pathway impact value. For more detailed information, such as the number of metabolites in the pathways (total number/hits/significant hits) and p-values, please refer to the supplementary information.

## Discussion

We performed integrated exposomics and metabolomics to detect the levels of exposure to contaminants and metabolite levels in cord serum. This comprehensive approach allowed us to assess the combined impact of environmental exposures and metabolic profiles on autoimmune diseases in the ABIS cohort. In our previous study we found similarities in metabolic profiles across different autoimmune diseases at birth (Hyotylainen et al. 2023). In order to avoid class imbalance problems, here we pooled all individual diseases together. We detected 20 contaminants, encompassing several PFAS compounds, Bisphenol S, and mycotoxins like Deoxynivalenol (DON), in cord blood samples from both control and case groups. Previous studies, including our own, have reported detectable levels of PFAS compounds (McGlinchey et al. 2020; Sinisalu et al. 2020), Bisphenol S exposure (Liu et al. 2017), and the presence of mycotoxins, including Deoxynivalenol (DON), in cord blood samples (Nielsen et al. 2011). These findings provide a backdrop for our investigation into the associations between these contaminants and autoimmune diseases in the ABIS cohort. We were able to demonstrate significant differences in the exposure levels of certain contaminants, such as Perfluorooctanoic acid Branched 2, Environmental Contaminant 1, Perfluorooctanoic acid Linear 1, Perfluorooctanoic acid Linear 2, and Methylparaben, in cord blood between the control and case groups. However, it is important to note that while these differences were statistically significant, the effect sizes were relatively modest. This suggests that while contaminants do play a role in distinguishing between controls and autoimmune diseases, they are unlikely to be the sole risk factors. Various factors including genetics, environmental triggers, and lifestyle factors, and their mutual interactions, contribute to the development of autoimmune diseases (Ellis et al. 2014; Oresic et al. 2013; Oresic et al. 2008; Rewers and Ludvigsson 2016; Sen et al. 2019; Sen et al. 2020; Sinisalu et al. 2020; Vermeulen et al. 2020; Virolainen et al. 2023).

Our study revealed differences in exposure and metabolite profiles between individuals who later developed autoimmune diseases and controls, particularly in relation to Z-score, mothers’ age, and BMI. This suggests that there may be differences in maternal factors between the two groups even at birth. We also observed that high levels of exposure to environmental contaminants were associated with changes in amino acid and free fatty acid profiles in the cord blood metabolome. Although we previously found a significant impact on lipid profiles, particularly triacylglycerols, the strength of association is weaker compared to the effect on polar metabolites (Hyotylainen et al. 2023). Among the 20 contaminants measured in our study, DON, Bisphenol S, and some branched PFAS compounds are the primary predictors of changes in cord serum metabolic profiles. While the associations between PFAS exposure and their marked effect on metabolism leading to autoimmune diseases have been well documented in previous studies (Ehrlich et al. 2023; McGlinchey et al. 2020; Rudzanova et al. 2023; Sinisalu et al. 2020), the exposure to DON and BPS and their impact on autoimmune diseases is less studied.

While our study detected Bisphenol S (BPS) and not Bisphenol A (BPA), it’s noteworthy that BPA, a common chemical found in plastics, has been associated with alterations in amino acid metabolism (Wang et al. 2018). BPA has been linked to changes in phenylalanine, tryptophan, tyrosine, lysine, and arginine metabolism, with a particular impact on female infants (Khan et al. 2017). In the case of BPS, it was shown to have sex- and diet-dependent effects on the development of type 1 diabetes (T1D) in non-obese diabetic (NOD) mice. Female mice exposed to BPS on a soy-based diet exhibited delayed T1D development, while males showed increased insulin resistance (Xu et al. 2019). These findings suggest that both BPA and BPS can influence metabolism and immune responses, potentially contributing to autoimmune diseases like T1D, although there is less evidence regarding the effect of BPS in humans.

Deoxynivalenol (DON) exposure in pregnant women has been reported in various studies. In the UK, pregnant women from diverse backgrounds showed detectable urinary DON levels, with South Asian women having higher exposure, primarily from bread consumption (Hepworth et al. 2012). Similarly, in Norway, DON, a common mycotoxin in cereals, was found in various cereal-based foods, potentially affecting the immune system, particularly in infants and young children (Sundheim et al. 2017). In pregnant Egyptian women, DON co-occurred with other mycotoxins, raising concerns about maternal and fetal health (Piekkola et al. 2012).

These findings emphasize the importance of assessing DON exposure in pregnant women and its potential health implications. DON exposure, prevalent in grains, adversely affects the immune system in both humans and animals and has been linked to alterations in gut microbiota (Liao et al. 2018). This immunotoxicity induced by DON involves mechanisms such as MAPK activation, ER stress, and mitochondrial signaling pathways (Liao et al. 2018).

To delve deeper into the potential mechanisms underlying these associations, we conducted pathway analysis of DON exposure on polar metabolites within both control and case groups. This analysis revealed that DON had distinct impacts on metabolic pathways in these groups. In the control group, DON exposure was associated with alterations in ‘Tyrosine and Tryptophan metabolism,’ indicating potential effects on amino acid pathways. Conversely, the case group showed associations between DON exposure and ‘Glutathione Metabolism,’ ‘Alanine and Aspartate Metabolism,’ and ‘Glycerophospholipid metabolism,’ suggesting disruptions in antioxidant defense systems and lipid metabolism. These findings align with previous research indicating that DON can induce oxidative stress by reducing antioxidant enzyme activity and enhancing lipid peroxidation (Mishra et al. 2014). The marked association between DON and phosphatidylcholines in our study suggests a potential link between mycotoxin exposure and alterations in lipid metabolism, particularly in the context of phosphatidylcholines. This finding is noteworthy, as specific phosphatidylcholines were previously identified as persistently down-regulated in children who later progressed to islet autoimmunity (Johnson et al. 2019) and clinical type 1 diabetes (T1D) (Oresic et al. 2008). Thus, the oxidative stress response and its impact on lipid metabolism, triggered by DON exposure, may play a pivotal role in the pathogenesis of autoimmune diseases, warranting further investigation.

These differential effects of DON exposure on metabolic pathways between control and case groups highlight the intricate relationship between environmental exposures, metabolism, and immune dysregulation in the context of autoimmune diseases. While our study contributes to our understanding of the metabolic consequences of DON exposure, it’s essential to consider these findings within the broader context of various factors, including genetics, environmental triggers, and lifestyle factors, which collectively contribute to the development of autoimmune diseases. Understanding these effects is crucial when assessing DON exposure during pregnancy and its potential health consequences. In this study, the median age of diagnosis of autoimmune diseases was higher compared to previous studies in genetically high-risk cohorts (Oresic et al. 2013; Oresic et al. 2008; Sen et al. 2019). Despite some common metabolic patterns (Hyotylainen et al. 2023), there were differences and limitations to consider. One important limitation of our study was the small sample size within each disease group, which restricted our analysis. Another limitation of our study was the lack of maternal exposure and longitudinal exposure data at different time points between birth and the onset of autoimmune diseases, which could explain their age-dependent progression.

Our previous studies in T1D (McGlinchey et al. 2020) and CD (Sinisalu et al. 2020) cohorts have mainly focus on the associations of PFAS exposure with the disease risk. Here we detected the levels of other contaminants such as Bisphenol S and some mycotoxins including DON and a-Zearalanol, which potentially show the differences in exposures. Mycotoxins are common contaminants of cereals and grains, and exposure to them is also associated with autoimmune disorders (Gayathri et al. 2018; Kraft et al. 2021; Liao et al. 2018; Rotter et al. 1996). This emphasizes the need for caution and control over mycotoxin exposure, particularly during pregnancy and critical developmental stages.

Altogether, our results show that high prenatal exposure to environmental contaminants associated with altered cord serum metabolite levels and may result in the progression of autoimmune diseases in the ABIS cohort. Other factors such as Z-score, maternal age and BMI are associated with contaminant exposure levels. Mechanistic studies are required to elucidate pathways of disease progression upon exposure.

## Author contributions

Bagavathy Shanmugam Karthikeyan: Formal analysis, Software, Visualization, Writing - Original Draft; Tuulia Hyötyläinen: Conceptualization, methodology, formal analysis, investigation, writing – original draft, supervision, project administration, funding acquisition; Tannaz Ghaffarzadegan: Writing – review and editing, methodology; Eric W. Triplett: Conceptualization, writing – review and editing; Matej Orešič: Conceptualization, writing – original draft, formal analysis, supervision; Johnny Ludvigsson: Conceptualization, writing – review and editing, funding acquisition, design and management of the ABIS cohort study.

## Acknowledgements

BSK thanks Partho Sen for his guidance in Methods and Alex Dickens for his insightful discussions. The authors thank Dirk Repsilber for his support in using the UCS-1 server from Örebro University for computations. This work was supported by the Swedish Research Council [grant number 2020-03674; to TH], Formas [grant number 2019-00869; to TH], and by the “Inflammation in human early life: targeting impacts on life-course health” (INITIALISE) consortium funded by the Horizon Europe Program of the European Union under Grant Agreement 101094099 (to T.H., E.W.T., M.O., and J.L.). The ABIS cohort study (JL) was supported by Barndiabetesfonden (Swedish Child Diabetes Foundation), the Swedish Council for Working Life and Social Research [grant numbers FAS2004-1775 and FAS2004–1775], Swedish Research Council [Grant/Award Numbers: K2005-72 × -11242-11A and K2008-69 × -20826-01-4, K2008-69 × -20826-01-4], Östgöta Brandstodsbolag, Medical Research Council of Southeast Sweden (FORSS), JDRF Wallenberg Foundation [Grant/Award Number: K 98-99D-12813-01A], Joanna Cocozza Foundation and ALF- and LfoU grants from Region Östergötland and Linköping University, Sweden.

## Data accessibility

Data are available upon reasonable request after ethical approvement and an appropriate institutional collaboration agreement. These data are not available to access in a repository owing to concern that the identity of patients might be revealed inadvertently.

## Supplementary Material

**Fig. S1.**
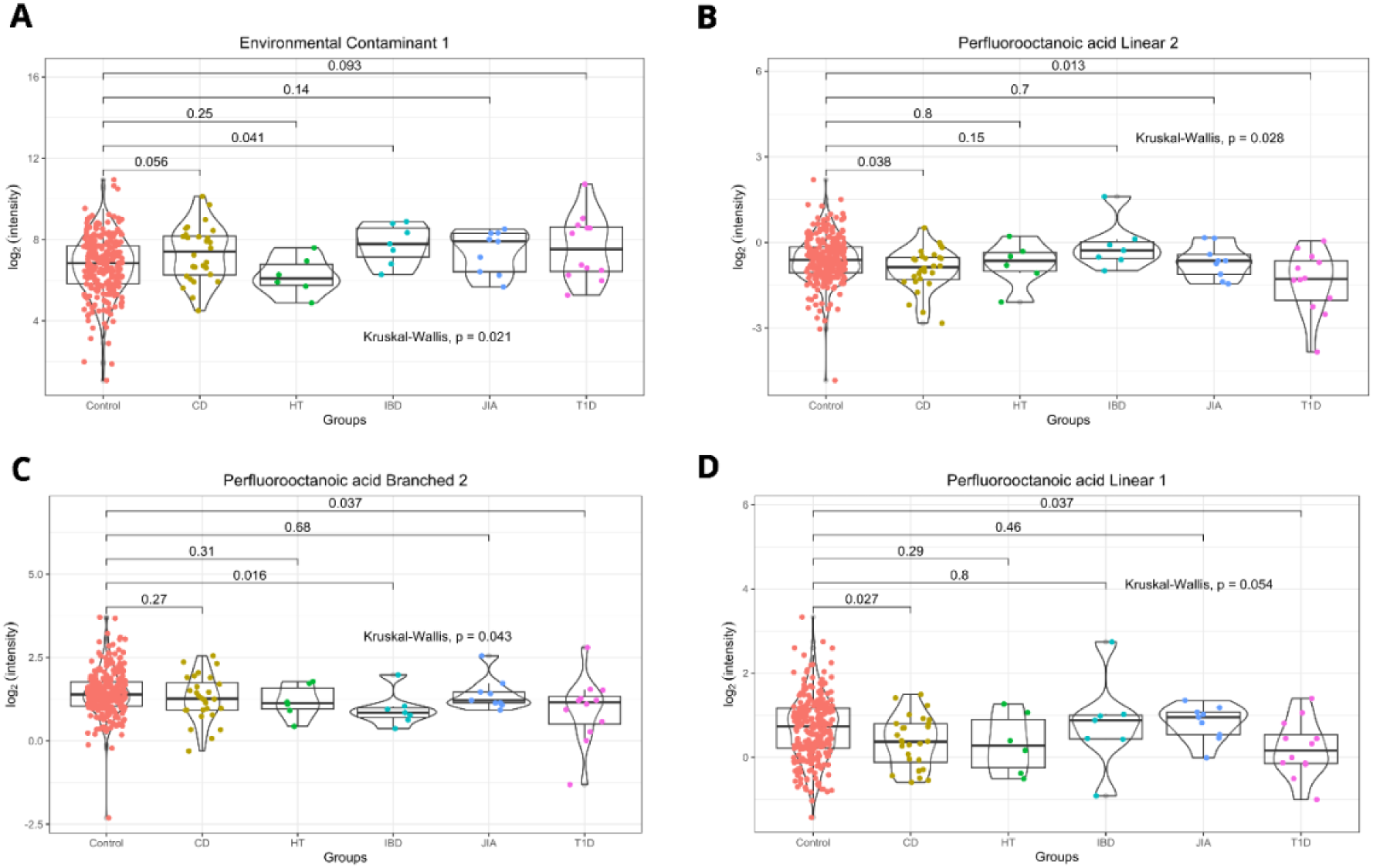
Box plots that show the levels of selected contaminants in the ABIS cohort at the individual disease levels. The violin plots (A-D) on top of the box plots illustrate the distribution of the selected contaminants (log2 intensities). To compare multiple group means, we used the Kruskal-Wallis Test, and for pairwise comparison against the reference (Control), we used the Wilcoxon Test. The p-values are provided to indicate the significance levels for the mean differences between the two groups (control vs. cases) for each contaminant (A-D). Specifically, p < 0.05 indicates statistical significance, and p < 0.1 suggests a trend toward significance. CD, HT, IBD, JIA, and T1D refer to Celiac disease, Hypothyroidism, Crohn’s disease, Juvenile Idiopathic Arthritis and Type 1 Diabetes respectively.

**Fig. S2.**
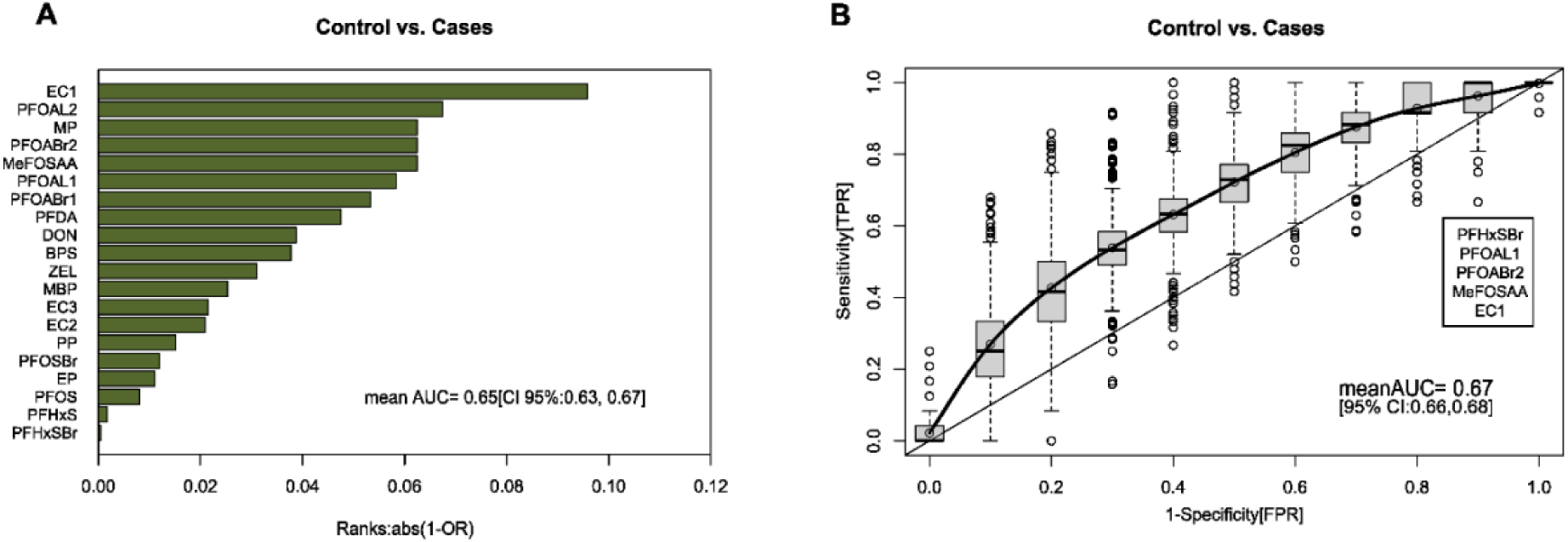
Classification of controls and immune-mediated diseases using contaminant exposure as predictors. In panel A, the ranks of the predictors (contaminants) obtained from the Logistic ridge regression (LRR) model, adjusted by Z-Score, Maternal age and BMI, are presented. The greatest contributing contaminants (predictors) that aided in the classification of control vs. cases (mean AUC = 0.65, 95% CI: 0.63–0.67) are shown at the top of the chart. In panel B, the Receiver Operating Characteristic (ROC) and AUC values from stepwise-predictive LRR models (10-fold cross-validation) are shown. An optimal set of five contaminants (predictors) (AUC = 0.67, 95% CI: 0.66–0.68) associated with the classification of control vs. cases are presented.

**Fig. S3.**
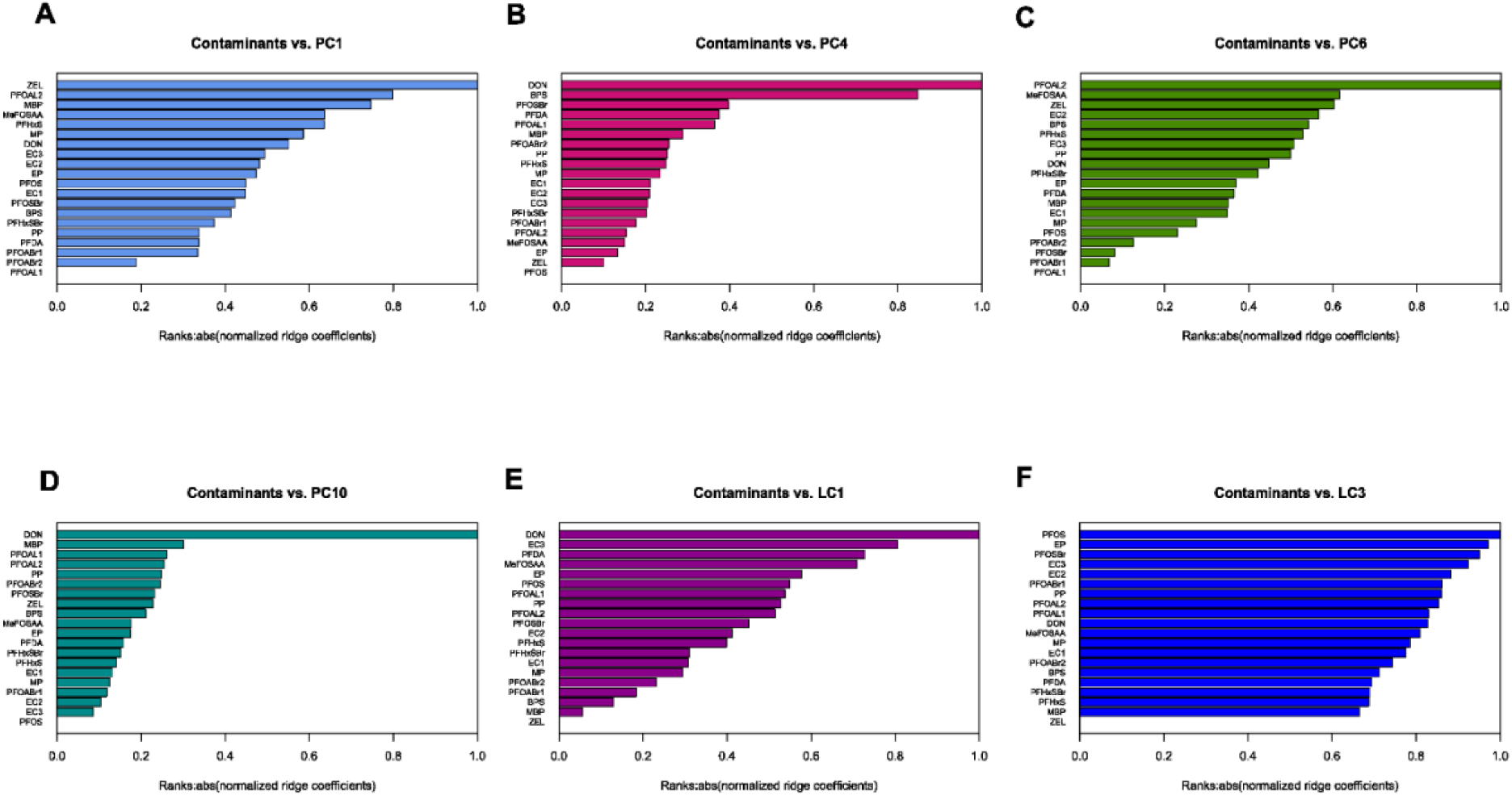
Association between exposure to environmental contaminants and the alteration of cord serum metabolites. The bar plots (A-F) show the linear predictors of changes in cord serum metabolite profiles, which are analysed in the form of metabolite clusters (Polar metabolite and lipid clusters). At the top of each bar plot (A-F), the most contributing contaminants (predictors) associated with selected metabolite clusters are shown. The ranks of the predictors are based on their absolute normalized (ridge) regression coefficients. The bar plots (A-D) represent the contaminant exposure as linear predictors of changes in polar metabolite clusters PC1, PC4, PC6 and PC10, while the bar plots (E-F) represent the contaminant exposure as linear predictors of changes in lipid clusters LC1 and LC3.

**Table S1.** List of contaminants, clusters and their level of identification based on the Metabolomics Standards Initiative (MSI).

| Full name | Abbreviation | Cluster | Level of identification |
| --- | --- | --- | --- |
| Bisphenol S | BPS | CC1 | Level 1 |
| Deoxynivalenol | DON | CC1 | Level 1 |
| Monobutyl phthalate | MBP | CC1 | Level 2 |
| Perfluorodecanoic acid | PFDA | CC1 | Level 1 |
| Methylperfluorooctane sulfonamidoacetic acid | MeFOSAA | CC1 | Level 2 |
| Environmental Contaminant 1 | EC1 | CC1 | Level 2 |
| Environmental Contaminant 2 | EC2 | CC1 | Level 2 |
| α-Zearalanol | ZEL | CC1 | Level 1 |
| Ethylparaben | EP | CC2 | Level 1 |
| Methylparaben | MP | CC2 | Level 1 |
| Propylparaben | PP | CC2 | Level 1 |
| Perfluorohexanesulfonic acid Branched | PFHxSBr | CC3 | Level 1 |
| Perfluorohexanesulfonic acid | PFHxS | CC3 | Level 1 |
| Perfluorooctanoic acid Branched 1 | PFOABr1 | CC4 | Level 1 |
| Perfluorooctanoic acid Linear 1 | PFOAL1 | CC4 | Level 1 |
| Perfluorooctanoic acid Branched 2 | PFOABr2 | CC4 | Level 1 |
| Perfluorooctanoic acid Linear 2 | PFOAL2 | CC4 | Level 1 |
| Perfluorooctanesulfonic acid Branched | PFOSBr | CC4 | Level 1 |
| Perfluorooctanesulfonic acid | PFOS | CC4 | Level 1 |
| Environmental Contaminant 3 | EC3 | CC4 | Level 2 |

**Table S2.** Two-way analysis of variance (ANOVA) for cord serum lipids (cluster LC1 to LC8) impacted by contaminants exposure (Contaminant cluster CC1 to CC4). The samples were grouped based on contaminants exposure quartiles (1 to 4) and Groups (Control/Cases). The statistical significance levels are represented by p-values (p < 0.05 marked bold, p-values < 0.1 in italics).

| Contaminant cluster | Lipid cluster | Factor 1: Contaminant quartiles | Factor 2: Groups (Control, Cases) | Interactions F1*F2 |
| --- | --- | --- | --- | --- |
| CC1 | LC1 | 0.291 | 0.203 | 0.467 |
| CC1 | LC2 | 0.271 | 0.674 | 0.568 |
| CC1 | LC3 | **$9.59 \times 10^{-4}$** | 0.884 | 0.701 |
| CC1 | LC4 | **$6.95 \times 10^{-4}$** | 0.983 | 0.850 |
| CC1 | LC5 | 0.519 | *0.055* | *0.064* |
| CC1 | LC6 | 0.265 | 0.124 | 0.111 |
| CC1 | LC7 | **$2.03 \times 10^{-2}$** | 0.919 | 0.936 |
| CC1 | LC8 | 0.869 | 0.449 | 0.260 |
| CC2 | LC1 | 0.706 | 0.195 | 0.895 |
| CC2 | LC2 | 0.683 | 0.719 | 0.471 |
| CC2 | LC3 | 0.814 | 0.901 | 0.992 |
| CC2 | LC4 | 0.531 | 0.973 | 0.958 |
| CC2 | LC5 | 0.995 | *0.071* | 0.947 |
| CC2 | LC6 | 0.731 | 0.176 | 0.608 |
| CC2 | LC7 | *0.096* | 0.960 | 0.139 |
| CC2 | LC8 | 0.298 | 0.378 | 0.917 |
| CC3 | LC1 | 0.578 | 0.220 | 0.721 |
| CC3 | LC2 | 0.283 | 0.803 | 0.856 |
| CC3 | LC3 | 0.276 | 0.981 | 0.730 |
| CC3 | LC4 | 0.581 | 0.840 | 0.747 |
| CC3 | LC5 | **$1.43 \times 10^{-4}$** | *0.061* | 0.433 |
| CC3 | LC6 | **$3.27 \times 10^{-3}$** | 0.194 | 0.329 |
| CC3 | LC7 | 0.909 | 0.967 | 0.160 |
| CC3 | LC8 | 0.179 | 0.455 | 0.919 |
| CC4 | LC1 | 0.249 | 0.141 | *0.074* |
| CC4 | LC2 | 0.246 | 0.534 | 0.416 |
| CC4 | LC3 | 0.194 | 0.713 | **0.027** |
| CC4 | LC4 | 0.221 | 0.868 | **0.032** |
| CC4 | LC5 | **0.019** | **0.026** | *0.068* |
| CC4 | LC6 | 0.361 | 0.123 | 0.348 |
| CC4 | LC7 | 0.172 | 0.774 | 0.521 |
| CC4 | LC8 | *0.095* | 0.296 | 0.523 |

**Table S3.** Post-Hoc test that followed a two-way analysis of variance for cord serum lipids (cluster LC1 to LC8) affected by contaminant exposure (Contaminant cluster CC1 to CC4). The Post-Hoc Tukeys’ HSD test was used for pairwise comparison between metabolite levels (along quartiles). The statistical significance levels are indicated by p-values, with values less than 0.05 marked in bold and values less than 0.1 in italics.

| Contaminant cluster | Lipid cluster | Factor 1: Contaminant quartiles |  |  |  |  |  |
| --- | --- | --- | --- | --- | --- | --- | --- |
|  |  | Q2-Q1 | Q3-Q1 | Q4-Q1 | Q3-Q2 | Q4-Q2 | Q4-Q3 |
| CC1 | LC1 | 1.000 | 0.379 | 0.762 | 0.371 | 0.755 | 0.925 |
| CC1 | LC2 | 0.407 | 0.820 | 0.271 | 0.905 | 0.994 | 0.785 |
| CC1 | LC3 | 0.317 | 0.106 | **$3.68 \times 10^{-4}$** | 0.942 | *0.091* | 0.289 |
| CC1 | LC4 | 0.166 | *0.081* | **$2.38 \times 10^{-4}$** | 0.988 | 0.156 | 0.292 |
| CC1 | LC5 | 0.991 | 0.904 | 0.866 | 0.980 | 0.711 | 0.467 |
| CC1 | LC6 | 0.216 | 0.659 | 0.911 | 0.864 | 0.585 | 0.962 |
| CC1 | LC7 | 0.759 | 0.154 | **0.018** | 0.675 | 0.204 | 0.837 |
| CC1 | LC8 | 0.988 | 0.870 | 0.915 | 0.971 | 0.987 | 0.999 |
| CC2 | LC1 | 0.929 | 0.952 | 0.992 | 0.665 | 0.988 | 0.849 |
| CC2 | LC2 | 0.957 | 0.633 | 0.864 | 0.903 | 0.993 | 0.977 |
| CC2 | LC3 | 0.986 | 0.935 | 0.992 | 0.788 | 0.926 | 0.990 |
| CC2 | LC4 | 0.993 | 0.694 | 0.943 | 0.523 | 0.839 | 0.952 |
| CC2 | LC5 | 1.000 | 0.998 | 0.998 | 0.997 | 0.997 | 1.000 |
| CC2 | LC6 | 0.968 | 0.798 | 0.745 | 0.968 | 0.946 | 1.000 |
| CC2 | LC7 | 0.148 | 0.989 | 0.325 | 0.277 | 0.975 | 0.516 |
| CC2 | LC8 | 0.995 | 0.341 | 0.640 | 0.483 | 0.786 | 0.961 |
| CC3 | LC1 | 0.609 | 0.625 | 0.862 | 1.000 | 0.971 | 0.975 |
| CC3 | LC2 | 0.807 | 0.220 | 0.914 | 0.731 | 0.995 | 0.585 |
| CC3 | LC3 | 0.870 | 0.943 | 0.729 | 0.997 | 0.277 | 0.384 |
| CC3 | LC4 | 1.000 | 0.985 | 0.756 | 0.977 | 0.787 | 0.539 |
| CC3 | LC5 | 0.216 | 0.343 | **$4.93 \times 10^{-5}$** | 0.994 | **0.048** | **0.024** |
| CC3 | LC6 | 0.490 | 0.273 | **0.001** | 0.979 | 0.101 | 0.231 |
| CC3 | LC7 | 0.999 | 0.983 | 0.986 | 0.953 | 0.998 | 0.893 |
| CC3 | LC8 | 0.299 | 0.702 | 0.174 | 0.910 | 0.991 | 0.772 |
| CC4 | LC1 | 0.969 | 0.655 | 0.629 | 0.376 | 0.353 | 1.000 |
| CC4 | LC2 | 0.759 | 0.826 | 0.178 | 0.999 | 0.718 | 0.646 |
| CC4 | LC3 | 0.863 | 0.462 | 0.167 | 0.903 | 0.573 | 0.932 |
| CC4 | LC4 | 0.812 | 0.727 | 0.157 | 0.999 | 0.620 | 0.718 |
| CC4 | LC5 | 0.984 | 0.215 | 0.139 | 0.101 | *0.059* | 0.996 |
| CC4 | LC6 | 0.999 | 0.881 | 0.470 | 0.818 | 0.389 | 0.895 |
| CC4 | LC7 | 0.993 | 0.504 | 0.449 | 0.345 | 0.300 | 1.000 |
| CC4 | LC8 | 0.928 | 0.754 | 0.315 | 0.379 | *0.093* | 0.886 |

**Table S4.** Two-way analysis of variance (ANOVA) for cord serum polar metabolites (cluster PC1 to PC12) impacted by contaminants exposure (Contaminant cluster CC1 to CC4). The samples were grouped based on contaminants exposure quartiles (1 to 4) and Groups (Control/Cases). The statistical significance levels are represented by p-values (p < 0.05 marked bold, p-values < 0.1 in italics).

| Contaminant cluster | Polar metabolite cluster | Factor 1: Contaminant quartiles | Factor 2: Groups (Control, Cases) | Interactions F1*F2 |
| --- | --- | --- | --- | --- |
| CC1 | PC1 | $<2 \times 10^{-16}$ | 0.685 | 0.331 |
| CC1 | PC2 | $<2 \times 10^{-16}$ | **0.022** | 0.516 |
| CC1 | PC3 | 0.780 | 0.237 | 0.380 |
| CC1 | PC4 | **$3.11 \times 10^{-11}$** | **0.001** | 0.876 |
| CC1 | PC5 | **0.026** | 0.554 | 0.401 |
| CC1 | PC6 | $<2 \times 10^{-16}$ | 0.338 | 0.378 |
| CC1 | PC7 | **0.017** | 0.726 | 0.533 |
| CC1 | PC8 | *0.082* | 0.963 | 0.533 |
| CC1 | PC9 | *0.085* | 0.703 | 0.824 |
| CC1 | PC10 | **$1.23 \times 10^{-11}$** | 0.967 | 0.731 |
| CC1 | PC11 | **$4.09 \times 10^{-9}$** | **$3.98 \times 10^{-4}$** | 0.544 |
| CC1 | PC12 | *0.068* | 0.356 | 0.824 |
| CC2 | PC1 | **0.025** | 0.574 | 0.186 |
| CC2 | PC2 | **$1.89 \times 10^{-4}$** | **0.097** | 0.817 |
| CC2 | PC3 | 0.836 | 0.250 | 0.723 |
| CC2 | PC4 | **0.021** | **0.004** | 0.637 |
| CC2 | PC5 | 0.338 | 0.498 | 0.789 |
| CC2 | PC6 | **0.001** | 0.236 | 0.446 |
| CC2 | PC7 | **0.006** | 0.735 | 0.122 |
| CC2 | PC8 | **0.006** | 0.631 | **0.091** |
| CC2 | PC9 | 0.718 | 0.577 | 0.250 |
| CC2 | PC10 | *0.051* | 0.727 | 0.567 |
| CC2 | PC11 | 0.469 | **0.004** | 0.793 |
| CC2 | PC12 | 0.162 | 0.310 | **0.011** |
| CC3 | PC1 | **$2.02 \times 10^{-6}$** | 0.589 | 0.829 |
| CC3 | PC2 | 0.131 | 0.199 | 0.550 |
| CC3 | PC3 | 0.115 | 0.259 | 0.665 |
| CC3 | PC4 | **0.044** | **0.011** | 0.515 |
| CC3 | PC5 | *0.095* | 0.538 | 0.396 |
| CC3 | PC6 | **0.017** | 0.160 | 0.273 |
| CC3 | PC7 | 0.509 | 0.738 | **0.004** |
| CC3 | PC8 | **0.002** | 0.809 | 0.628 |
| CC3 | PC9 | 0.905 | 0.591 | 0.232 |
| CC3 | PC10 | *0.094* | 0.554 | 0.953 |
| CC3 | PC11 | **0.044** | **0.005** | 0.384 |
| CC3 | PC12 | 0.822 | 0.318 | 0.148 |
| CC4 | PC1 | 0.242 | 0.468 | 0.861 |
| CC4 | PC2 | 0.376 | 0.118 | 0.678 |
| CC4 | PC3 | **0.034** | 0.143 | 0.855 |
| CC4 | PC4 | 0.138 | **0.002** | 0.666 |
| CC4 | PC5 | **0.027** | 0.273 | 0.473 |
| CC4 | PC6 | **0.001** | 0.398 | 0.617 |
| CC4 | PC7 | 0.686 | 0.833 | **0.032** |
| CC4 | PC8 | **0.010** | 0.427 | **0.090** |
| CC4 | PC9 | 0.659 | 0.738 | 0.525 |
| CC4 | PC10 | **0.019** | 0.621 | 0.943 |
| CC4 | PC11 | 0.183 | **0.002** | 0.677 |
| CC4 | PC12 | 0.579 | 0.401 | **0.006** |

**Table S5.**
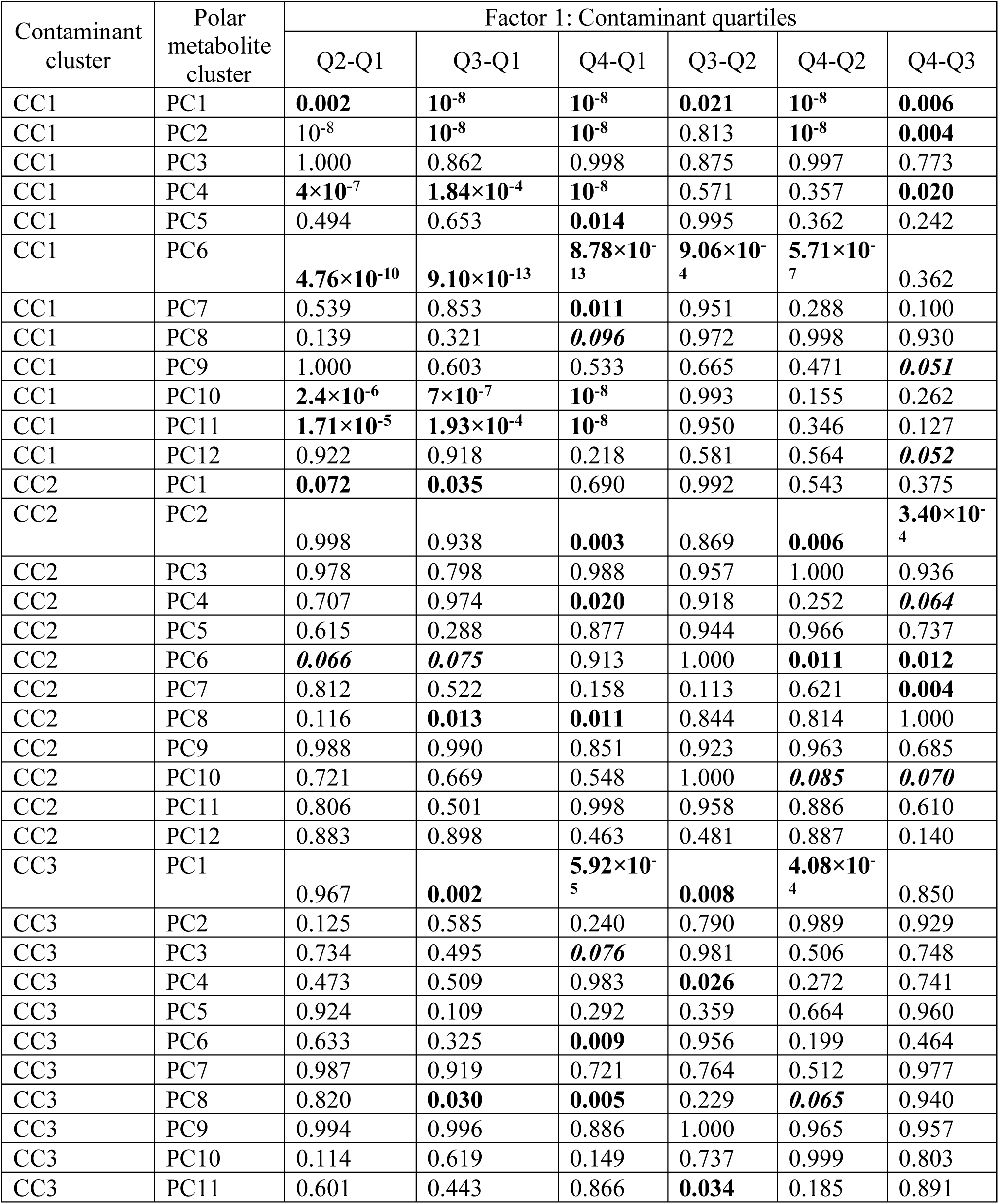
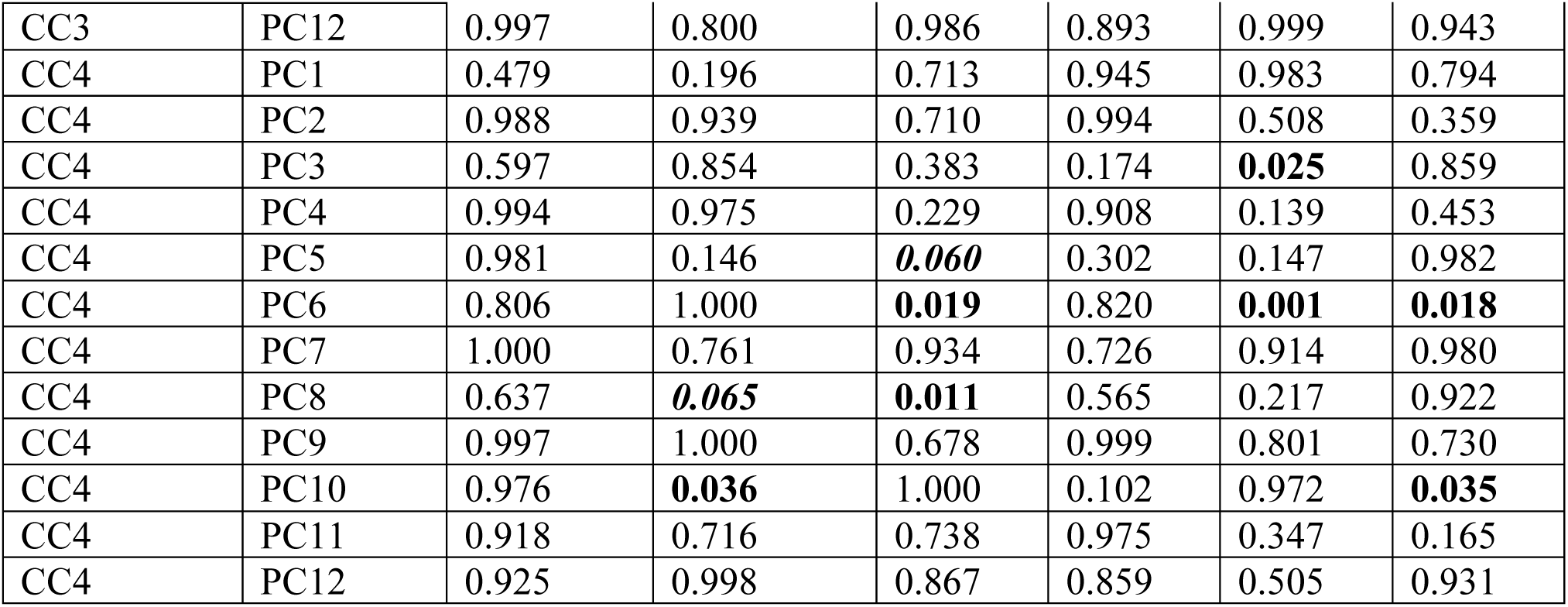
Post-Hoc test that followed a two-way analysis of variance for cord serum polar metabolites (cluster PC1 to PC12) affected by contaminant exposure (Contaminant cluster CC1 to CC4). The Post-Hoc Tukeys’ HSD test was used for pairwise comparison between metabolite levels (along quartiles). The statistical significance levels are indicated by p-values, with values less than 0.05 marked in bold and values less than 0.1 in italics.

**Table S6.**
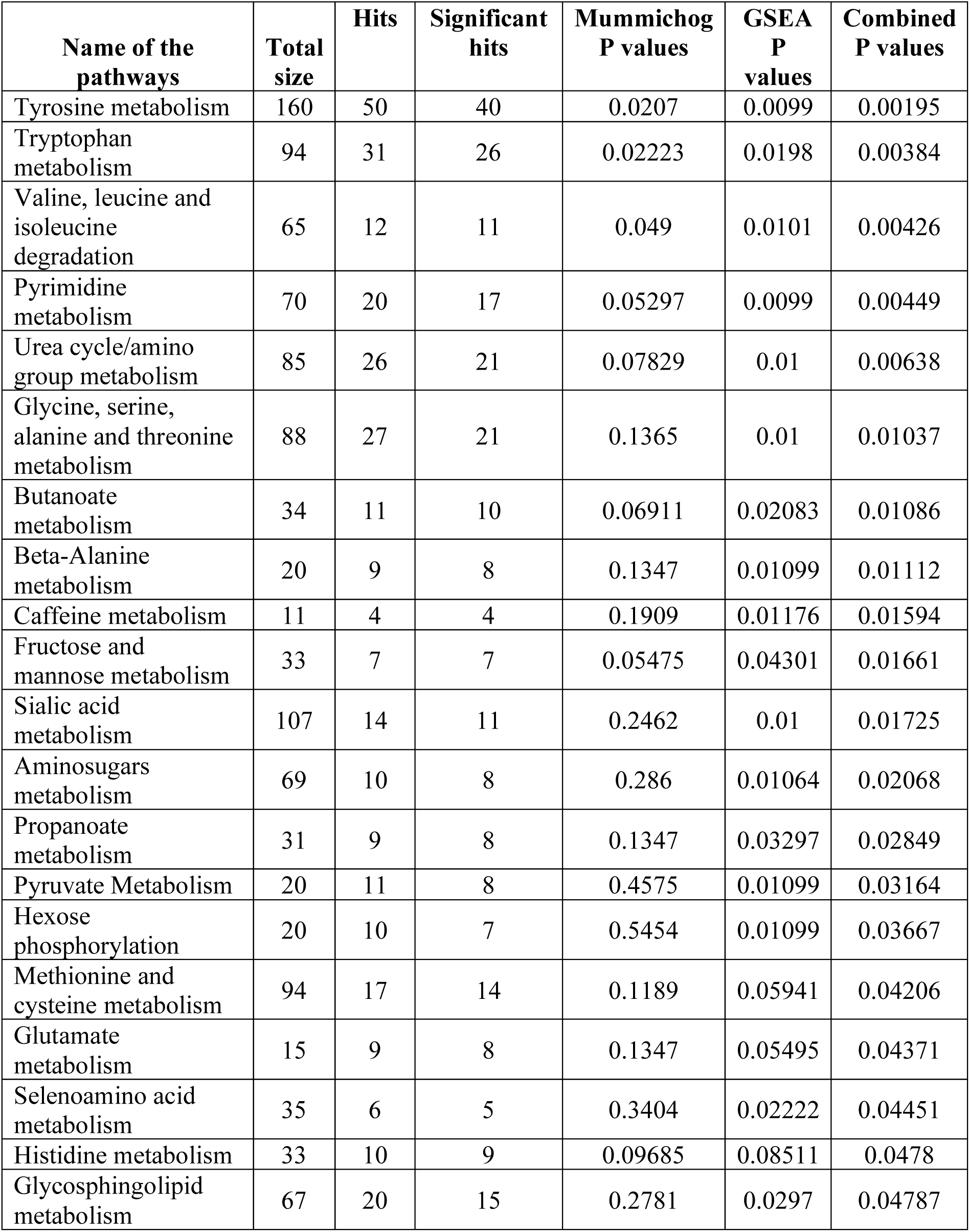
provides information on the pathways identified through pathway enrichment analysis using MFN pathway maps for controls. It includes the pathways, their corresponding p-values, and the number of metabolites in each pathway, including the total size, hits, and significant hits. The combined p-value was calculated by combining GSEA and Mummichog scores. This table lists only those pathways that have a combined p-value of less than 0.05.

**Table S7.**
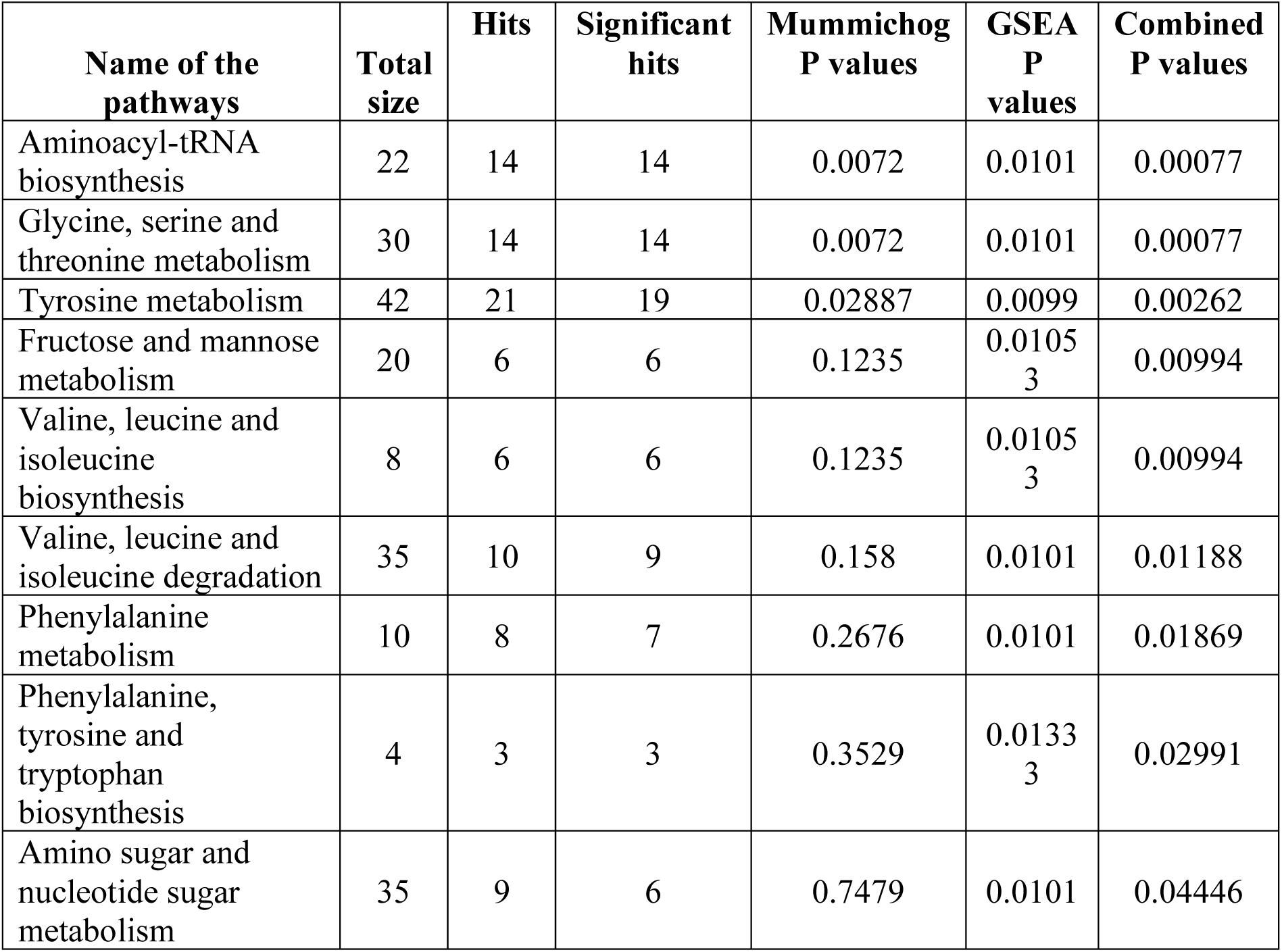
provides information on the pathways identified through pathway enrichment analysis using KEGG pathway maps for controls. It includes the pathways, their corresponding p-values, and the number of metabolites in each pathway, including the total size, hits, and significant hits. The combined p-value was calculated by combining GSEA and Mummichog scores. This table lists only those pathways that have a combined p-value of less than 0.05.

**Table S8.**
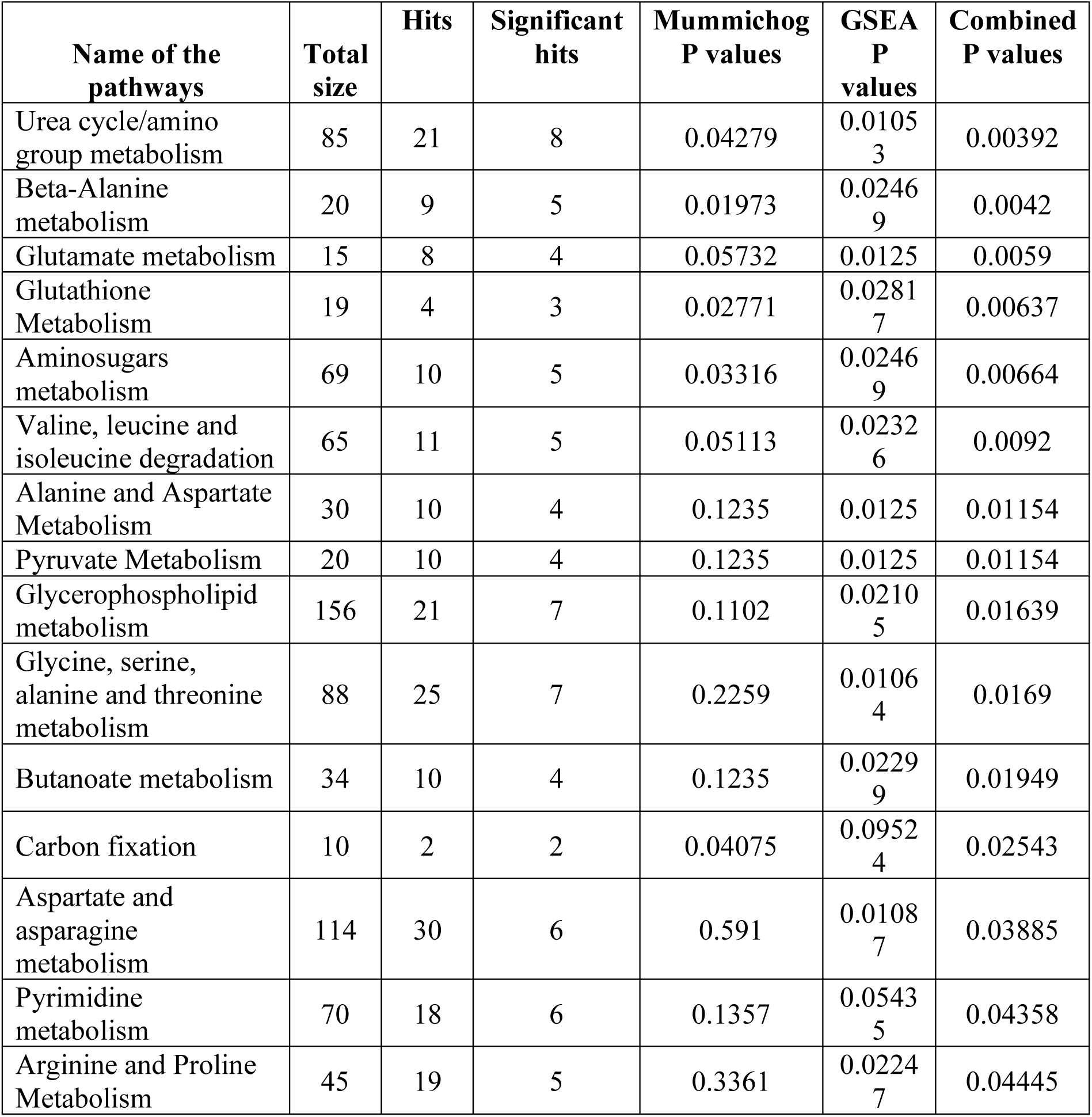
provides information on the pathways identified through pathway enrichment analysis using MFN pathway maps for cases. It includes the pathways, their corresponding p-values, and the number of metabolites in each pathway, including the total size, hits, and significant hits. The combined p-value was calculated by combining GSEA and Mummichog scores. This table lists only those pathways that have a combined p-value of less than 0.05.

**Table S9.**
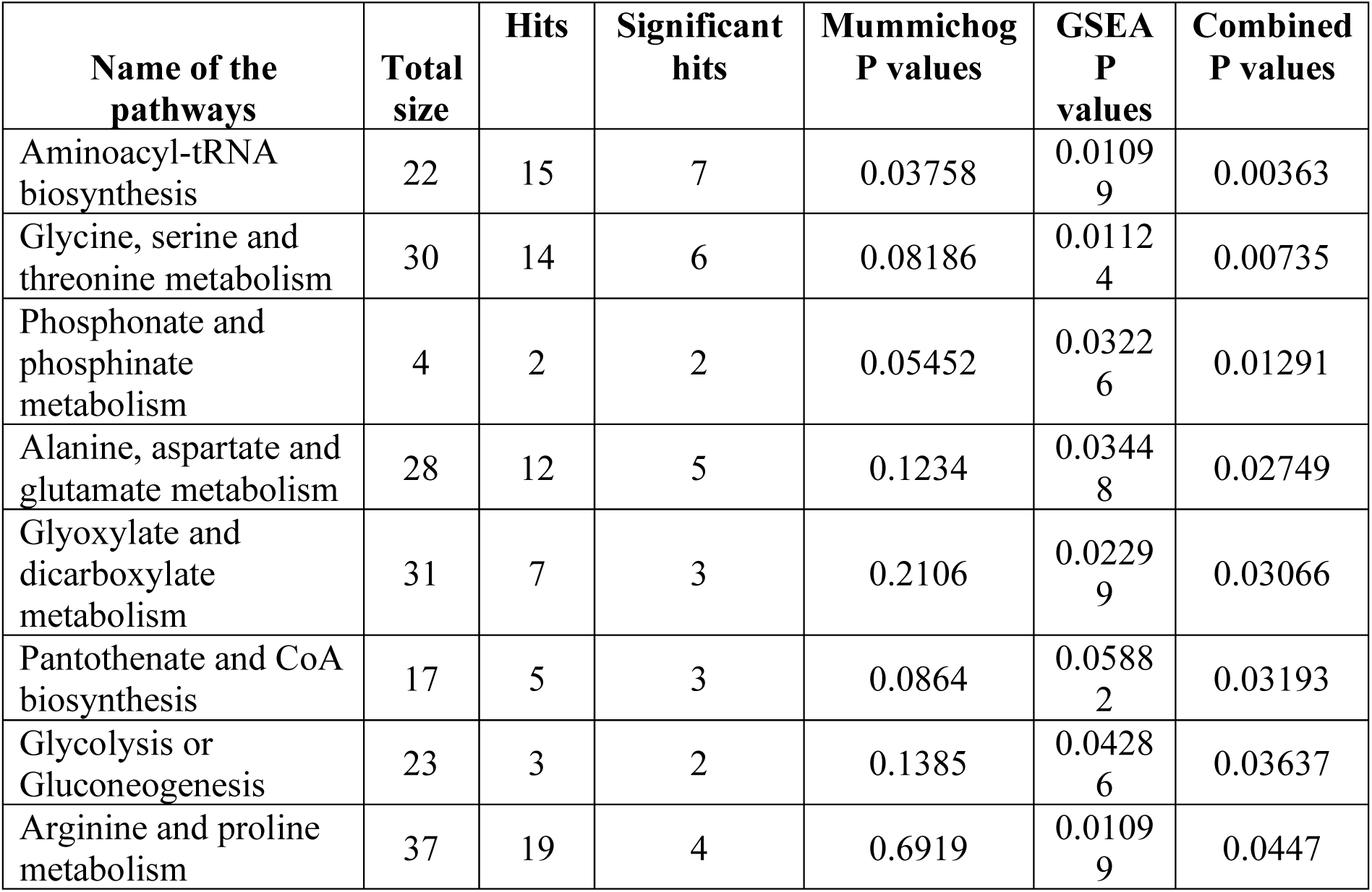
provides information on the pathways identified through pathway enrichment analysis using KEGG pathway maps for cases. It includes the pathways, their corresponding p-values, and the number of metabolites in each pathway, including the total size, hits, and significant hits. The combined p-value was calculated by combining GSEA and Mummichog scores. This table lists only those pathways that have a combined p-value of less than 0.05.

